# Transcriptomic Gene Network Profiling and Weak Signal Detection for Prediction of Ovarian Cancer Occurrence, Survival, and Severity by Integrating Bulk and Single-cell RNAseq Data

**DOI:** 10.1101/2023.12.21.23300414

**Authors:** Yanming Li, Mihaela Sardiu, Devin C. Koestler, Fengwei Yang, Md Tamzid Islam, Stephan Komladzei, Murshalina Akhter

## Abstract

**Background:** Ovarian cancer (OC) is a significant gynecological malignancy characterized by its high mortality rate, poor long-term survival rate, and late-stage diagnosis. OC is the 5th leading cause of cancer death among woman and counts 2.1% of all cancer death. OC survival rates are much lower than other cancers that affect woman. Its 5-year survival rate is less than 50%. Only ∼17% of OC patients are diagnosed within the early stage. The majority are diagnosed at an advanced stage, making early detection and effective treatment critical challenges. Currently, the identified OC predictive genes are still very sparse, resulting in pool prognostic performance. There exists unmet needs to identify novel prognostic gene biomarkers for OC occurrence, survival, and clinical stages to promote the likelihood of survival and to perform optimal treatments or therapeutic strategies at the earliest stage possible.

**Methods:** Previous RNAseq analysis on OC focused on detecting differentially expressed (DE) genes only. Many genes, although having weak marginal differential effects, may still exude strong predictive effects on disease outcomes though regulating other DE genes. In this work, we employed a new machine learning method, netLDA, to detect such predictive coregulating genes with weak marginal DE effects for predicting OC occurrence, 5-year survival, and clinical stage. The netLDA detects predictive gene networks (PGN) containing strong DE genes as hub genes and detects coregulating weak genes within the PGNs. The network structures of the detected PGNs along with the strong and weak genes therein are then used in outcome prediction on test datasets.

**Results:** We identified different sets of signature genes for OC occurrence, survival, and clinical stage. Previously identified prognostic genes, such as *EPCAM, UBE2C, CHD1L, TP53*,*CD24*, *WFDC2*, and *FANCI,* were confirmed. We also identified novel predictive coregulating weak genes including *GIGYF2, GNPAT, RAD54L*, and *ELL.* Many of the detected predictive gene networks and coregulating weak genes therein overlapped with OC-related biological pathways such as KEGG *tight junction, ribosome*, and *cell cycle* pathways. The detection and incorporation of the gene networks and weak genes significantly improved the prediction performance. Cellular mapping of selected feature genes using single-cell RNAseq data further revealed the heterogeneous expression distributions of the signature genes on different cell types.

**Conclusions:** We established a transcriptomic gene network profile for OC prediction. The novel genes detected provide new targets for early diagnostics and new drug development for OC.

## Introduction

Ovarian cancer (OC) is a significant gynecological malignancy characterized by its high incidence rate, poor survival, late-stage diagnosis, and limited treatment options [1,2]. It ranks as one of the most lethal cancers and the 5th leading cause of cancer death among woman [3]. It counts 2.1% of all cancer death. Ovarian cancer survival rates are much lower than other cancers that affect woman. Its 5-year survival rate is less than 50%. Its survivors often face physical and psychological challenges, including long-term side effects of treatment, infertility, and anxiety about cancer recurrence. Women diagnosed before the cancer has spread have a much higher five-year survival rate than those diagnosed at a later stage. However, only ∼17% of ovarian cancer patients are diagnosed within the early stage. The majority of OC cases are diagnosed at an advanced stage, making early detection and effective treatment critical challenges [4].

Advances in transcriptomic and genomic research have provided new insights in the discovery of OC oncogenes. Dozens of OC susceptibility genes, such as *BRCA1* [5–8], *BRCA2* [5–7], *EPCAM* [9,10]*, TP53* [11,12]and *CHD1L* [13–15], have been identified over the past decades. However, the sensitivity and specificity using only these genes in prognostics remain suboptimal. Efforts of using machine learning (ML) methods to identify novel gene biomarkers for predicting OC occurrence are still ongoing.

Compared to cancer occurrence, less research in the literature has been conducted on prediction of OC survival [15,16] and clinical stages [17,18]. It is particularly of scientific interest to investigate whether it is the same or different sets of genes that contribute to OC development and progression (including survival and clinical stages). Identification of novel oncogenes for predicting OC patients’ survival and clinical stages would be extremely helpful to promote the likelihood of survival and optimal treatment or therapeutic strategies at the earliest stage as possible, even after the patients being diagnosed of OC.

Through a series of work, Li et al. [19–23] have shown that in cancer-genomic studies, some genes, even though having weak marginal differential effects (DE), may still exude strong prediction effects on disease outcomes though regulating other strong DE genes. These weak DE genes (or weak genes), together with their coregulated strong genes and the coregulations between them, form predictive gene networks (PDN). Detecting such PDNs and the weak genes therein and integrating them into disease outcome prediction could significantly improve the prediction accuracy. In this project, we used a novel cancer-genomics analytical tool that we recently developed: netLDA – network-based linear discriminant analysis (https://github.com/lyqglyqg/netLDA) – to detect predictive gene networks and strong/weak signature genes for predicting OC occurrence, 5-year survival, and clinical stages, using both bulk and single-cell RNAseq (scRNAseq) data.

By looking at the gene-gene coregulation networks and weak genes, novel signature genes were identified, and the outcome prediction accuracy was significantly increased. The results helped with a better understanding of the underlining dynamic mechanisms of OC development and progression. They may shed light on promotion of precision medicine and new gene therapy development.

## Materials

### Data acquisition and processing

Bulk RNAseq and clinical data of 419 OC patients from The Cancer Genome Atlas (TCGA) program and 88 non-disease controls from The Genotype-Tissue Expression (GTEx) project were combined and used as the training data for OC occurrence prediction. Bulk RNAseq data in GSE18521 for 53 OC tumor samples and 10 normal ovary tissue samples from the Gene Expression Omnibus (GEO) database were used as an independent test dataset in the case-control study. There were 11,069 mapped genes on both training and test datasets.

Bulk RNAseq and survival data from GSE26712 for 195 OC patients were downloaded from GEO and used as the training dataset in the survival prediction. The same types of data for 53 OC patients were downloaded from GSE18521 and used as a test dataset. There were 12,645 common genes mapped on both datasets. The reason for not using TCGA data as the training data is that TCGA subjects cross a wide range of OC stages, which are heavily confounded with the survival. We did not find a GEO dataset that contains both survival and clinical stage outcomes. Therefore, we used two GEO datasets (both of which contained only late-staged OC patients) as the training and test datasets to alleviate the confounding effect from clinical stages.

Bulk RNAseq and clinical data from 419 TCGA OC patients and from 77 GSE63885 OC patients were used as the training and test data, respectively, in the OC clinical stage prediction. There were 17,490 mapped genes on both training and test datasets.

Single-cell RNAseq data of 22,153 cells and 47,913 transcripts from GSE229343 were used in the scRNAseqs data analysis and cellular mapping for the signature genes selected in each study.

The RNAseq data went through quality control before analysis using R package edgeR [24] or Seurat [25]. For the bulk RNAseq data, genes with counts less than 10 for more than 70% of the samples were removed from analyses. For the scRNAseq data, cells with UMI numbers below 500, gene numbers below 300 or greater than 6,000, or mitochondrial-derived UMI counts of more than 15% were considered low-quality and were removed [103].

## Methods

Three studies were conducted for prediction of different OC outcomes: i) occurrence prediction of OC v.s. healthy, ii) 5-year survival prediction of survival longer than 5 years v.s. shorter than 5 years, and iii) severity prediction of clinical stage ≤ III v.s. V. The following methods were used in each study.

### PGN and network-based weak gene selections

We use the netLDA [20] in both feature selection and outcome prediction. **Figure 1** depicts the major steps of netLDA. First, netLDA selects top strong DE gene as hub genes according to their marginal DE effects. Then for each strong DE gene, netLDA selects its coregulated gene network containing its highly correlated genes (having a Pearson correlation coefficient 𝜌 with |𝜌| > 0.8). Next, netLDA assigns the following predictive score, or network-adjusted DE effect, to each gene in a selected coregulating network, 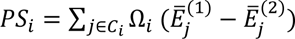, where 𝑖 and 𝑗 are gene indices, 𝐶_i_ is the set of genes connected to gene 𝑖 through a coregulation path, Ω_i_ is the precision matrix (inverse of the covariance matrix) that characterizes the coregulation information (directions and strengths) between genes in 𝐶_i_, and 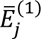 and 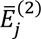 are the average expression level of gene 𝑗 in outcome groups 1 and 2, respectively. The predictive score integrates, for each targeted gene, how many other genes it coregulates, the strengths and directions of those regulations, and expression levels of its coregulated genes, as well as expression levels of the targeted gene itself. The most predictive genes are selected according to the strengths of their predictive scores.

**Figure 1.**
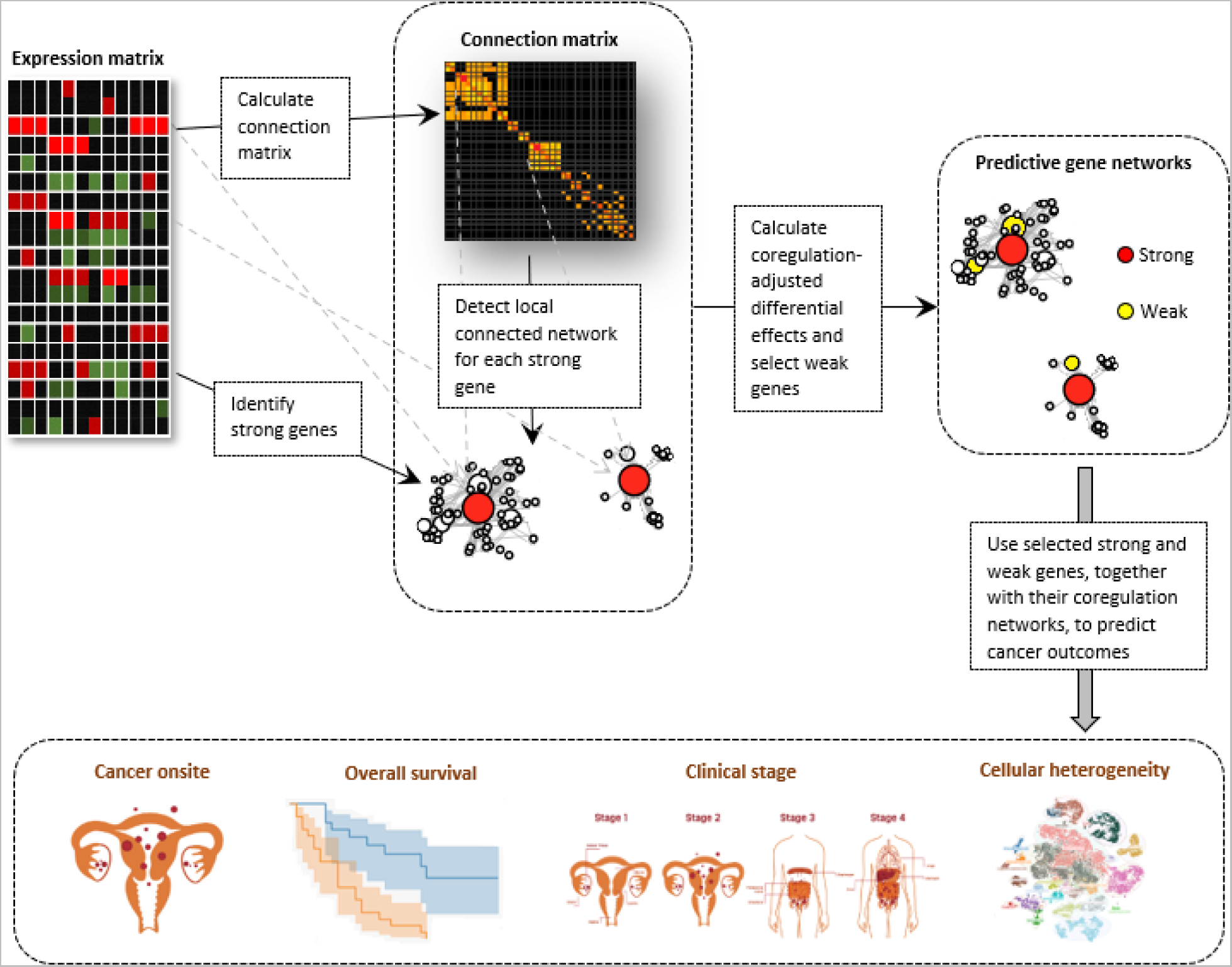
Analysis steps of netLDA. Step 1, netLDA selects top strong DE gene as hub coregulating genes. Step 2, for each strong DE gene, netLDA selects its coregulated gene network containing its highly correlated genes. Step 3, netLDA assigns the following predictive score to each gene in the coregulating network. The most predictive genes are selected according to the strength of their predictive scores, including strong DE genes and weak coregulating genes. Step 4, netLDA uses the selected strong and weak genes and their network structures to predict outcomes on the test data.

Selected predictive genes with small marginal DE effects are weak coregulating genes. For prediction, netLDA uses only the selected predictive genes and the coregulation network structures between them to predict outcomes on the test data. **Figure 2** explains the calculation of the predictive scores using toy example data. We also developed permutation tests to evaluate the significance of selected individual genes and PGNs.

**Figure 2.**
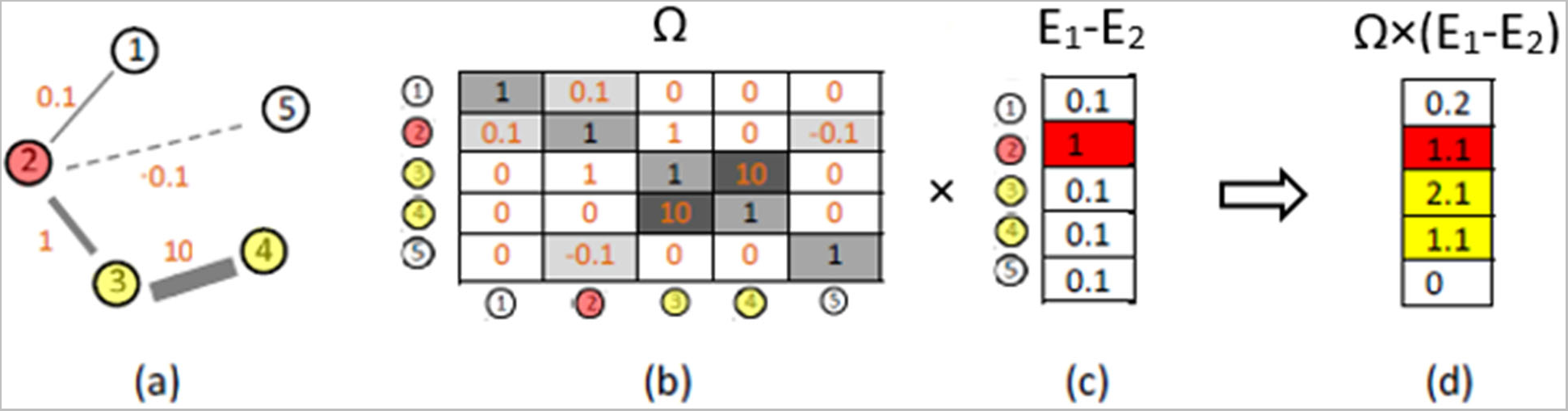
Detecting weakly differentiating genes but predictive by integrating gene-gene coregulations. **(a)** An example of coregulatory gene network containing five genes. Gene 2 has a strong marginally differentiating effect. The other four genes do not exhibit differentiating effects marginally, as shown in **(c)**. The unidirectional coregulations between the five genes are presented by the edge connections between them. Solid edges represent a positive coregulation and dashed edges represent a negative coregulation. If two genes are not connected by an edge, it means there is no known coregulation between them. The coregulation strength is represented by the width of the edges with the actual values marked aside. **(b)** Connection matrix corresponding to the network in (a). Numbers on the diagonal are the marginal variances for the five genes. Numbers off diagonal are the coregulation strength between genes. **(c)** Marginal expression differences between two groups. Only gene 2 exhibits a strong differential effect. **(d)** The coregulation adjusted differential effects are achieved by multiply the marginal effects by the connection matrix. Genes 3 and 4 also exhibit strong differential effects after the adjustment because of their coregulations with the strong gene 2.

### Prediction performance comparison with competing ML methods

We compared the prediction performance of netLDA with other commonly used cancer-genomics ML methods including Lasso [26], Ridge [27], ElasticNet [28], XGboost [29], and linear discriminate analysis (LDA) using only the strong genes and ignoring the coregulatory network structures between them. Prediction sensitivities, specificities, and the areas under the receiver operating characteristic curves (ROC) were evaluated to assess the prediction.

### Kepler-Meijer analysis

Kepler-Meijer (KM) analysis is a commonly used biomarker validation approach in cancer genomics studies. It compares the survival or KM curves between high- and low- expressed groups of a DE gene [30]. Here we generated KM curves according to the long- and short-term survival groups predicted by using the selected strong and weak genes, and their PGN structures. We compared the KM curves to the ones generated from only using the top strong genes’ expression levels.

### Gene set enrichment analysis

To validate our identified genes and PGNs from a biological perspective, we conducted gene set enrichment analysis (GSEA) [31], a knowledge-based approach for interpreting transcriptome profiles, using GeneOntology (GO) [32] and Kyoto Encyclopedia of Genes and Genomes (KEGG) [33] pathways. The selected strong/weak genes and PGNs were mapped to the top enriched KEGG pathways to confirm their oncological functionals.

### Cellular mapping for the selected genes

To reveal the cellular expression heterogeneity of the selected signature genes, we also performed a scRNAseq analysis for cell type profiling and cellular mapping of the selected genes.

## Results

### Predictive gene network and network-based gene selections

Top selected genes in the three studies are listed in **Table 1**. Marginal expression patterns for the selected genes are depicted in **Figure 3**. Top selected PGNs harboring the selected genes were listed in **Table 2**. Topological structures (illustrating the connection topologies) and connection matrices (illustrating the connection strengths) of the PNGs, along with marginal and network-adjusted

**Figure 3.**
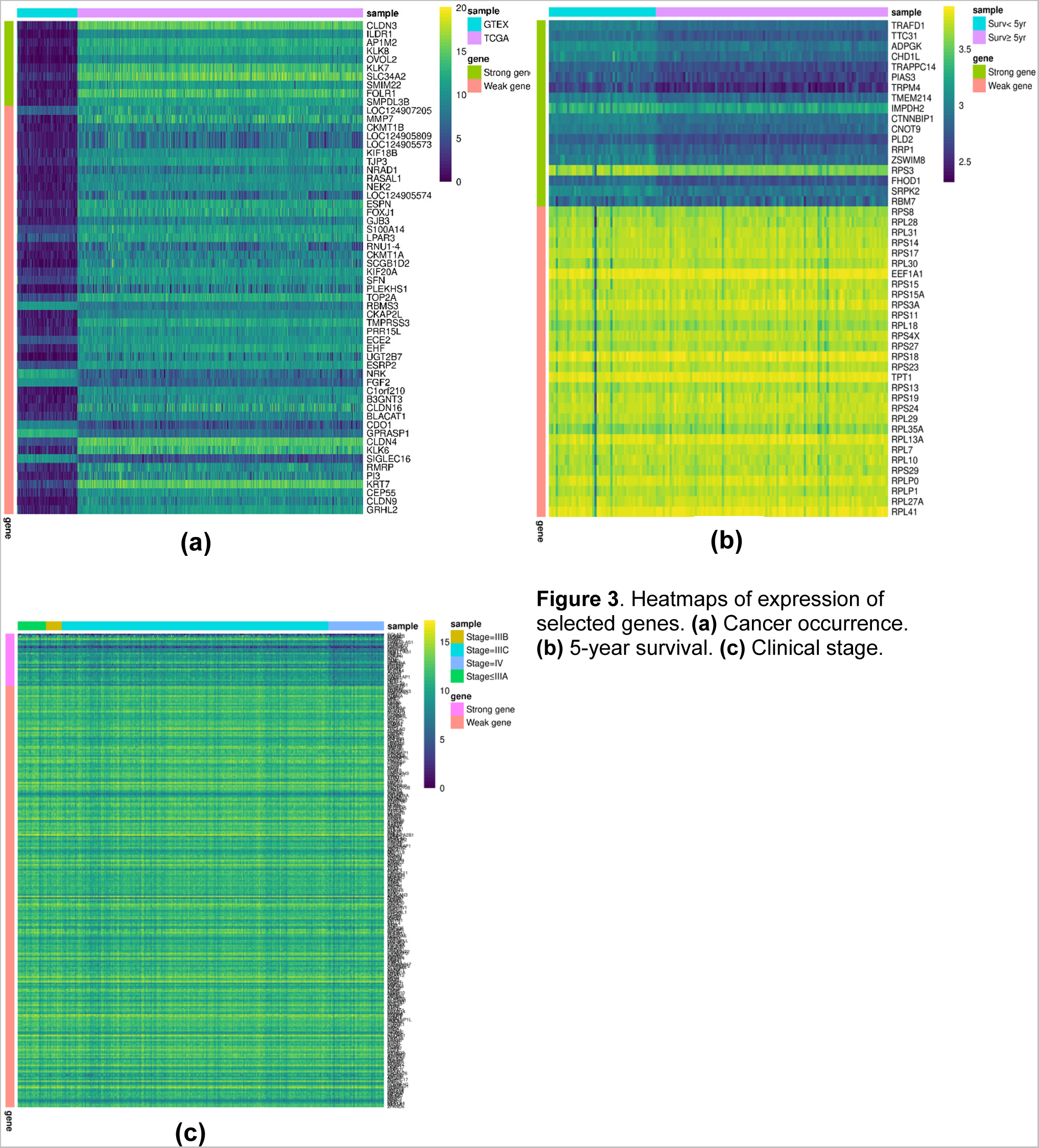
Heatmaps of expression of selected genes. **(a)** Cancer occurrence. **(b)** 5-year survival. **(c)** Clinical stage.

**Table 1.**
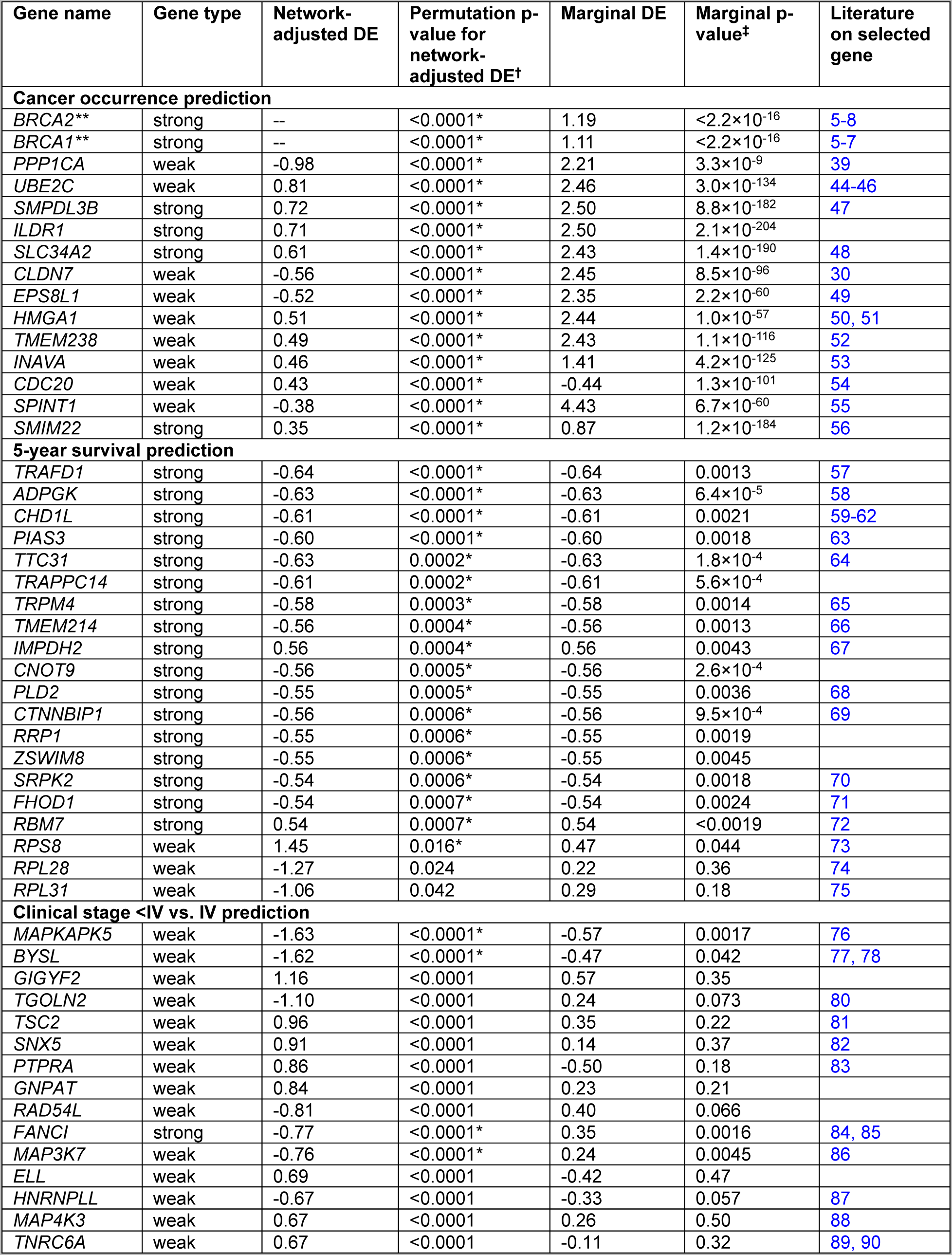
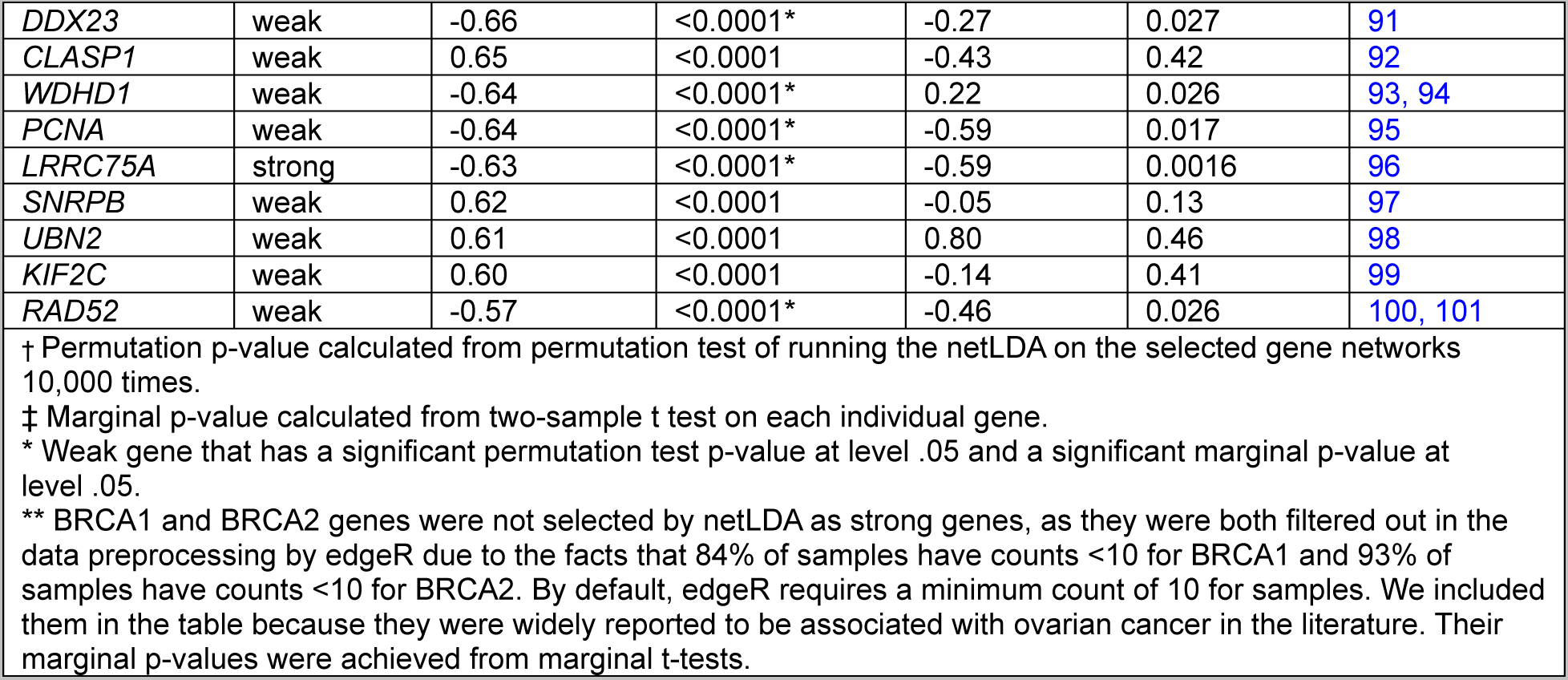
Top selected genes.

| Gene name | Gene type | Network-adjusted DE | Permutation p-value for network-adjusted DE<sup>†</sup> | Marginal DE | Marginal p-value<sup>‡</sup> | Literature on selected gene |
| --- | --- | --- | --- | --- | --- | --- |
| **Cancer occurrence prediction** |  |  |  |  |  |  |
| *BRCA2* ** | strong | -- | <0.0001* | 1.19 | <2.2×10<sup>-16</sup> | 5-8 |
| *BRCA1* ** | strong | -- | <0.0001* | 1.11 | <2.2×10<sup>-16</sup> | 5-7 |
| *PPP1CA* | weak | -0.98 | <0.0001* | 2.21 | 3.3×10<sup>-9</sup> | 39 |
| *UBE2C* | weak | 0.81 | <0.0001* | 2.46 | 3.0×10<sup>-134</sup> | 44-46 |
| *SMPDL3B* | strong | 0.72 | <0.0001* | 2.50 | 8.8×10<sup>-182</sup> | 47 |
| *ILDR1* | strong | 0.71 | <0.0001* | 2.50 | 2.1×10<sup>-204</sup> |  |
| *SLC34A2* | strong | 0.61 | <0.0001* | 2.43 | 1.4×10<sup>-190</sup> | 48 |
| *CLDN7* | weak | -0.56 | <0.0001* | 2.45 | 8.5×10<sup>-96</sup> | 30 |
| *EPS8L1* | weak | -0.52 | <0.0001* | 2.35 | 2.2×10<sup>-60</sup> | 49 |
| *HMGA1* | weak | 0.51 | <0.0001* | 2.44 | 1.0×10<sup>-57</sup> | 50, 51 |
| *TMEM238* | weak | 0.49 | <0.0001* | 2.43 | 1.1×10<sup>-116</sup> | 52 |
| *INAVA* | weak | 0.46 | <0.0001* | 1.41 | 4.2×10<sup>-125</sup> | 53 |
| *CDC20* | weak | 0.43 | <0.0001* | -0.44 | 1.3×10<sup>-101</sup> | 54 |
| *SPINT1* | weak | -0.38 | <0.0001* | 4.43 | 6.7×10<sup>-60</sup> | 55 |
| *SMIM22* | strong | 0.35 | <0.0001* | 0.87 | 1.2×10<sup>-184</sup> | 56 |
| **5-year survival prediction** |  |  |  |  |  |  |
| *TRAFD1* | strong | -0.64 | <0.0001* | -0.64 | 0.0013 | 57 |
| *ADPGK* | strong | -0.63 | <0.0001* | -0.63 | 6.4×10<sup>-5</sup> | 58 |
| *CHD1L* | strong | -0.61 | <0.0001* | -0.61 | 0.0021 | 59-62 |
| *PIAS3* | strong | -0.60 | <0.0001* | -0.60 | 0.0018 | 63 |
| *TTC31* | strong | -0.63 | 0.0002* | -0.63 | 1.8×10<sup>-4</sup> | 64 |
| *TRAPPC14* | strong | -0.61 | 0.0002* | -0.61 | 5.6×10<sup>-4</sup> |  |
| *TRPM4* | strong | -0.58 | 0.0003* | -0.58 | 0.0014 | 65 |
| *TMEM214* | strong | -0.56 | 0.0004* | -0.56 | 0.0013 | 66 |
| *IMPDH2* | strong | 0.56 | 0.0004* | 0.56 | 0.0043 | 67 |
| *CNOT9* | strong | -0.56 | 0.0005* | -0.56 | 2.6×10<sup>-4</sup> |  |
| *PLD2* | strong | -0.55 | 0.0005* | -0.55 | 0.0036 | 68 |
| *CTNNBIP1* | strong | -0.56 | 0.0006* | -0.56 | 9.5×10<sup>-4</sup> | 69 |
| *RRP1* | strong | -0.55 | 0.0006* | -0.55 | 0.0019 |  |
| *ZSWIM8* | strong | -0.55 | 0.0006* | -0.55 | 0.0045 |  |
| *SRPK2* | strong | -0.54 | 0.0006* | -0.54 | 0.0018 | 70 |
| *FHOD1* | strong | -0.54 | 0.0007* | -0.54 | 0.0024 | 71 |
| *RBM7* | strong | 0.54 | 0.0007* | 0.54 | <0.0019 | 72 |
| *RPS8* | weak | 1.45 | 0.016* | 0.47 | 0.044 | 73 |
| *RPL28* | weak | -1.27 | 0.024 | 0.22 | 0.36 | 74 |
| *RPL31* | weak | -1.06 | 0.042 | 0.29 | 0.18 | 75 |
| **Clinical stage <IV vs. IV prediction** |  |  |  |  |  |  |
| *MAPKAPK5* | weak | -1.63 | <0.0001* | -0.57 | 0.0017 | 76 |
| *BYSL* | weak | -1.62 | <0.0001* | -0.47 | 0.042 | 77, 78 |
| *GIGYF2* | weak | 1.16 | <0.0001 | 0.57 | 0.35 |  |
| *TGOLN2* | weak | -1.10 | <0.0001 | 0.24 | 0.073 | 80 |
| *TSC2* | weak | 0.96 | <0.0001 | 0.35 | 0.22 | 81 |
| *SNX5* | weak | 0.91 | <0.0001 | 0.14 | 0.37 | 82 |
| *PTPRA* | weak | 0.86 | <0.0001 | -0.50 | 0.18 | 83 |
| *GNPAT* | weak | 0.84 | <0.0001 | 0.23 | 0.21 |  |
| *RAD54L* | weak | -0.81 | <0.0001 | 0.40 | 0.066 |  |
| *FANCI* | strong | -0.77 | <0.0001* | 0.35 | 0.0016 | 84, 85 |
| *MAP3K7* | weak | -0.76 | <0.0001* | 0.24 | 0.0045 | 86 |
| *ELL* | weak | 0.69 | <0.0001 | -0.42 | 0.47 |  |
| *HNRNPLL* | weak | -0.67 | <0.0001 | -0.33 | 0.057 | 87 |
| *MAP4K3* | weak | 0.67 | <0.0001 | 0.26 | 0.50 | 88 |
| *TNRC6A* | weak | 0.67 | <0.0001 | -0.11 | 0.32 | 89, 90 |
| *DDX23* | weak | -0.66 | <0.0001* | -0.27 | 0.027 | 91 |
| *CLASP1* | weak | 0.65 | <0.0001 | -0.43 | 0.42 | 92 |
| *WDHD1* | weak | -0.64 | <0.0001* | 0.22 | 0.026 | 93, 94 |
| *PCNA* | weak | -0.64 | <0.0001* | -0.59 | 0.017 | 95 |
| *LRRC75A* | strong | -0.63 | <0.0001* | -0.59 | 0.0016 | 96 |
| *SNRPB* | weak | 0.62 | <0.0001 | -0.05 | 0.13 | 97 |
| *UBN2* | weak | 0.61 | <0.0001 | 0.80 | 0.46 | 98 |
| *KIF2C* | weak | 0.60 | <0.0001 | -0.14 | 0.41 | 99 |
| *RAD52* | weak | -0.57 | <0.0001* | -0.46 | 0.026 | 100, 101 |

† Permutation p-value calculated from permutation test of running the netLDA on the selected gene networks 10,000 times.
‡ Marginal p-value calculated from two-sample t test on each individual gene.
\* Weak gene that has a significant permutation test p-value at level .05 and a significant marginal p-value at level .05.
\*\* BRCA1 and BRCA2 genes were not selected by netLDA as strong genes, as they were both filtered out in the data preprocessing by edgeR due to the facts that 84% of samples have counts <10 for BRCA1 and 93% of samples have counts <10 for BRCA2. By default, edgeR requires a minimum count of 10 for samples. We included them in the table because they were widely reported to be associated with ovarian cancer in the literature. Their marginal p-values were achieved from marginal t-tests.

**Table 2.**
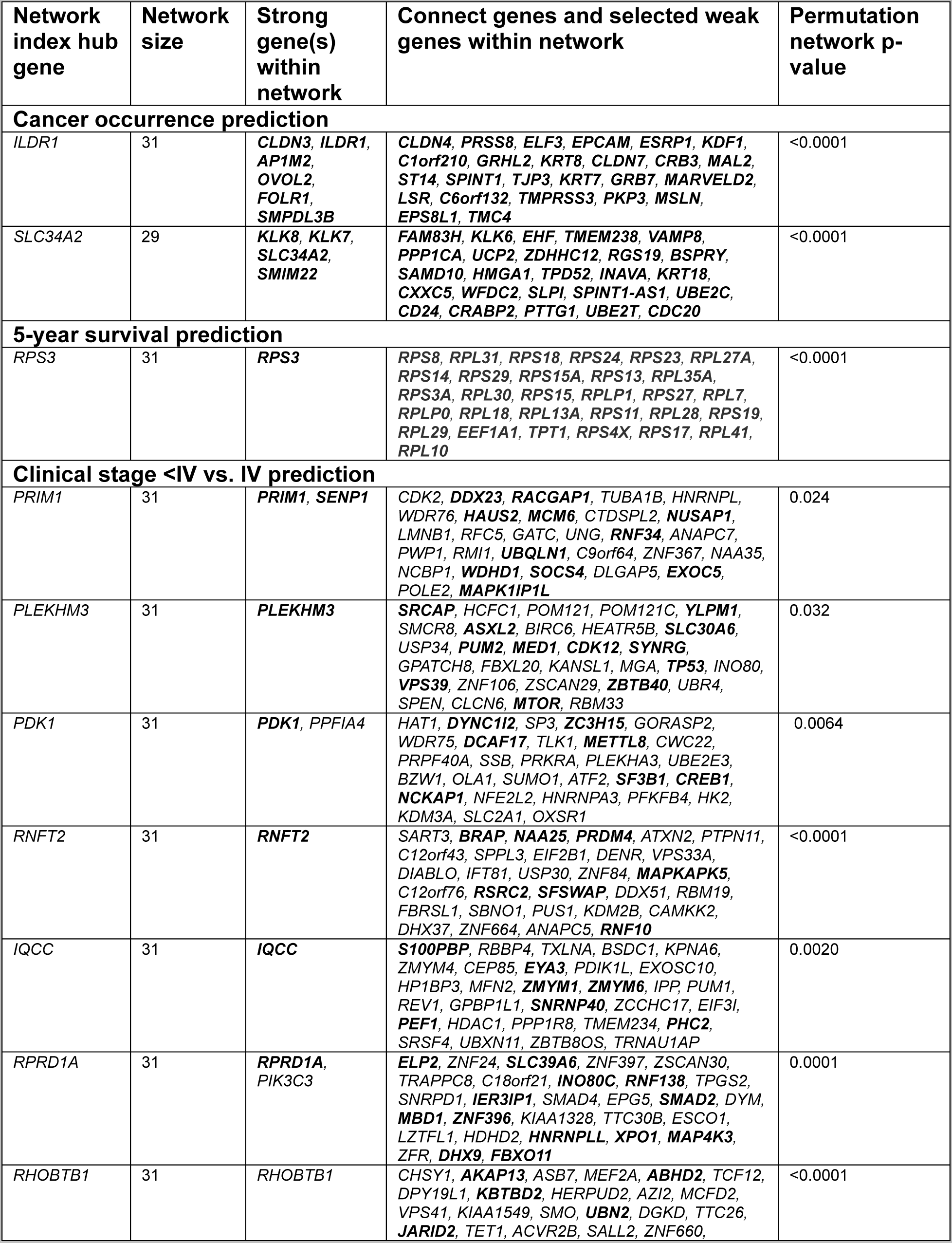
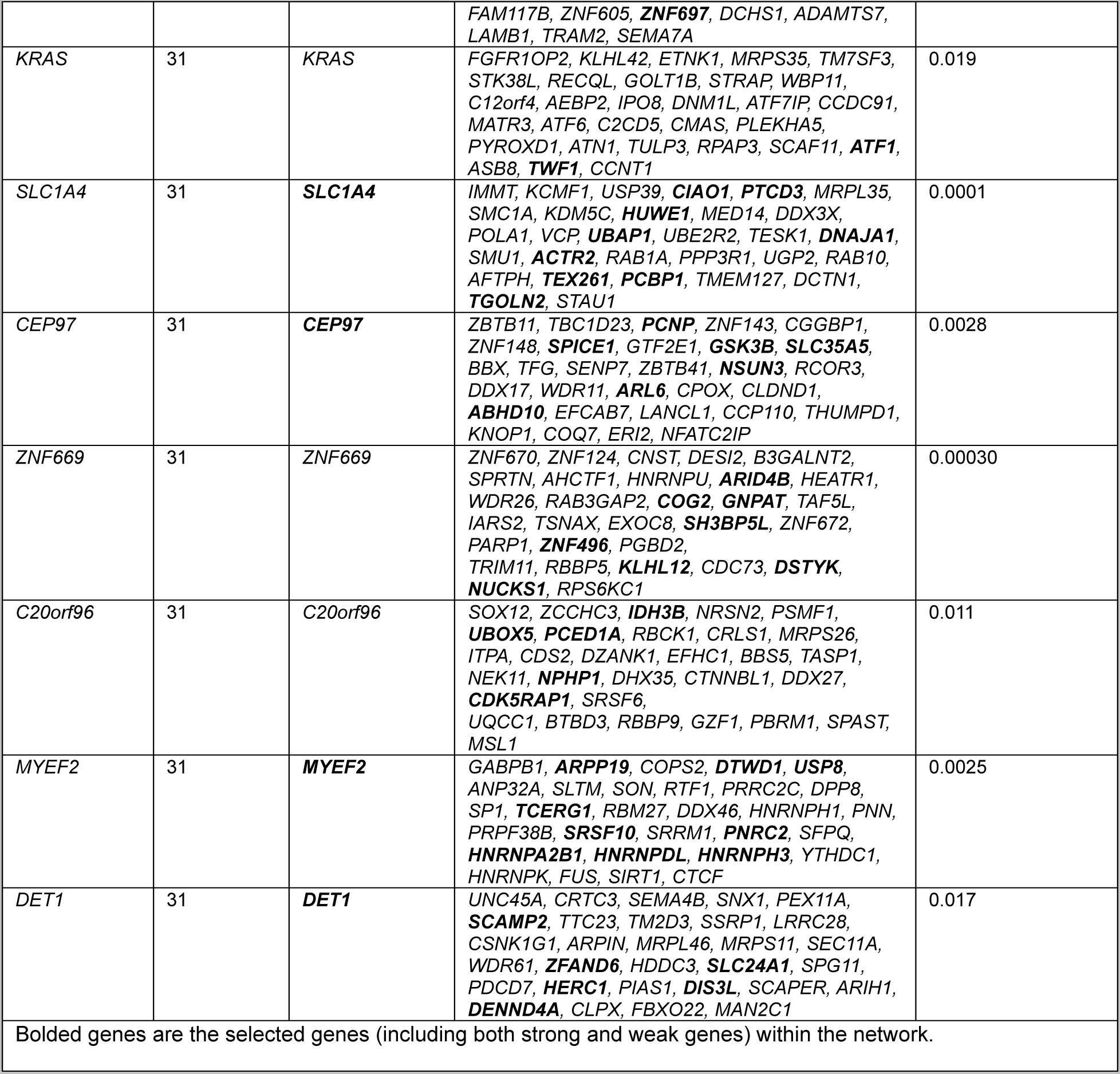
Top selected predictive gene networks.

DE effects of the genes within, were depicted in **Figure 4**. Most of the selected strong genes have both a significant marginal p-value (from marginal tests) and a significant permutation test p-value (<10^-4^, from a network-based test). While majority of the selected weak genes have only a significant permutation test p-value. This demonstrates that integrating of coregulation between genes helps to promote the significance of weak genes in their empirical distributions. For many of the top selected genes, we found literature evidence supporting their associations with OC (last column in **Table 1**). Most of the selected predictive gene networks have significant permutation test p-values (<0.05). Full lists of the selected genes and PGNs are given in the Supplemental Materials.

**Figure 4.**
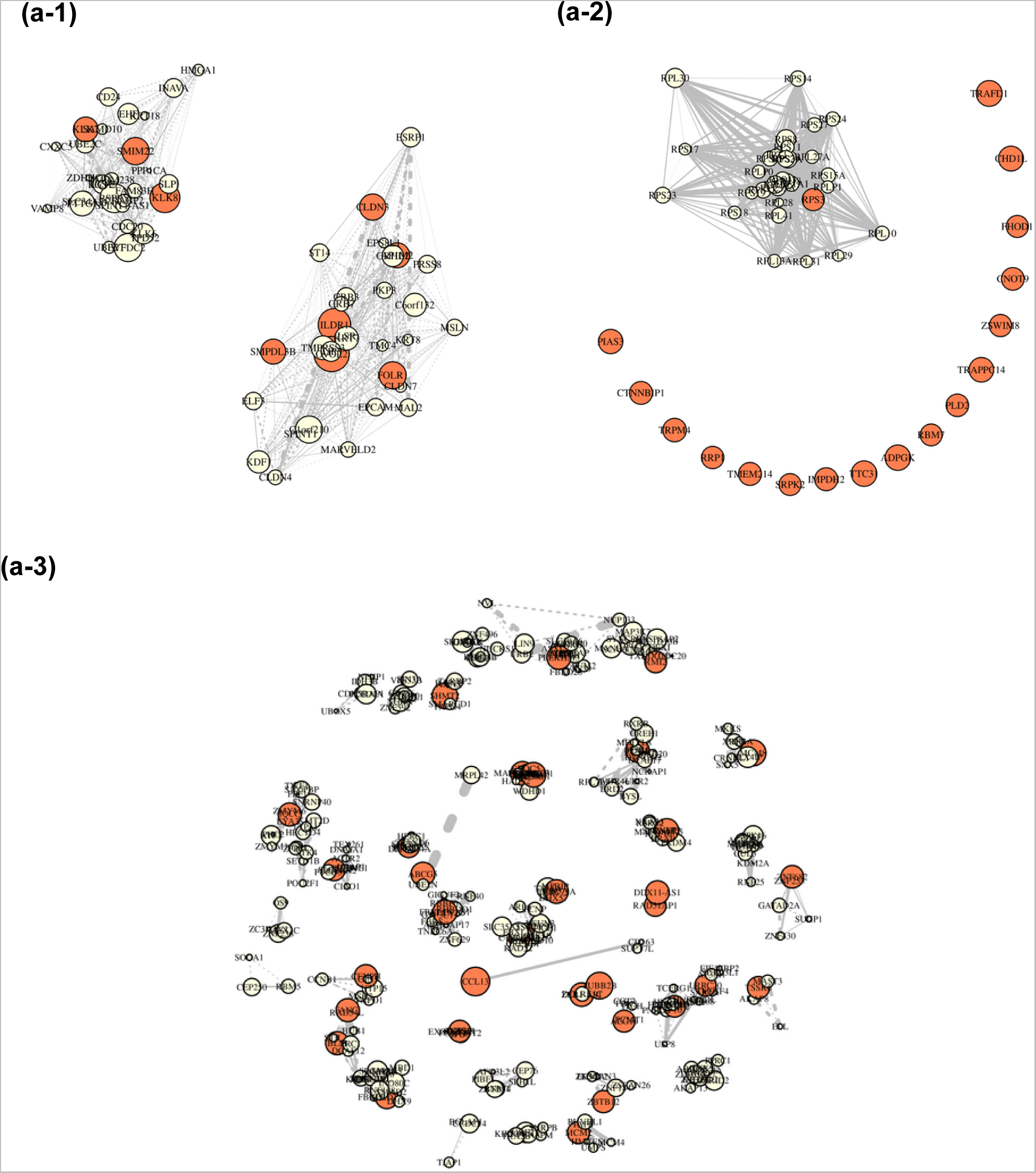
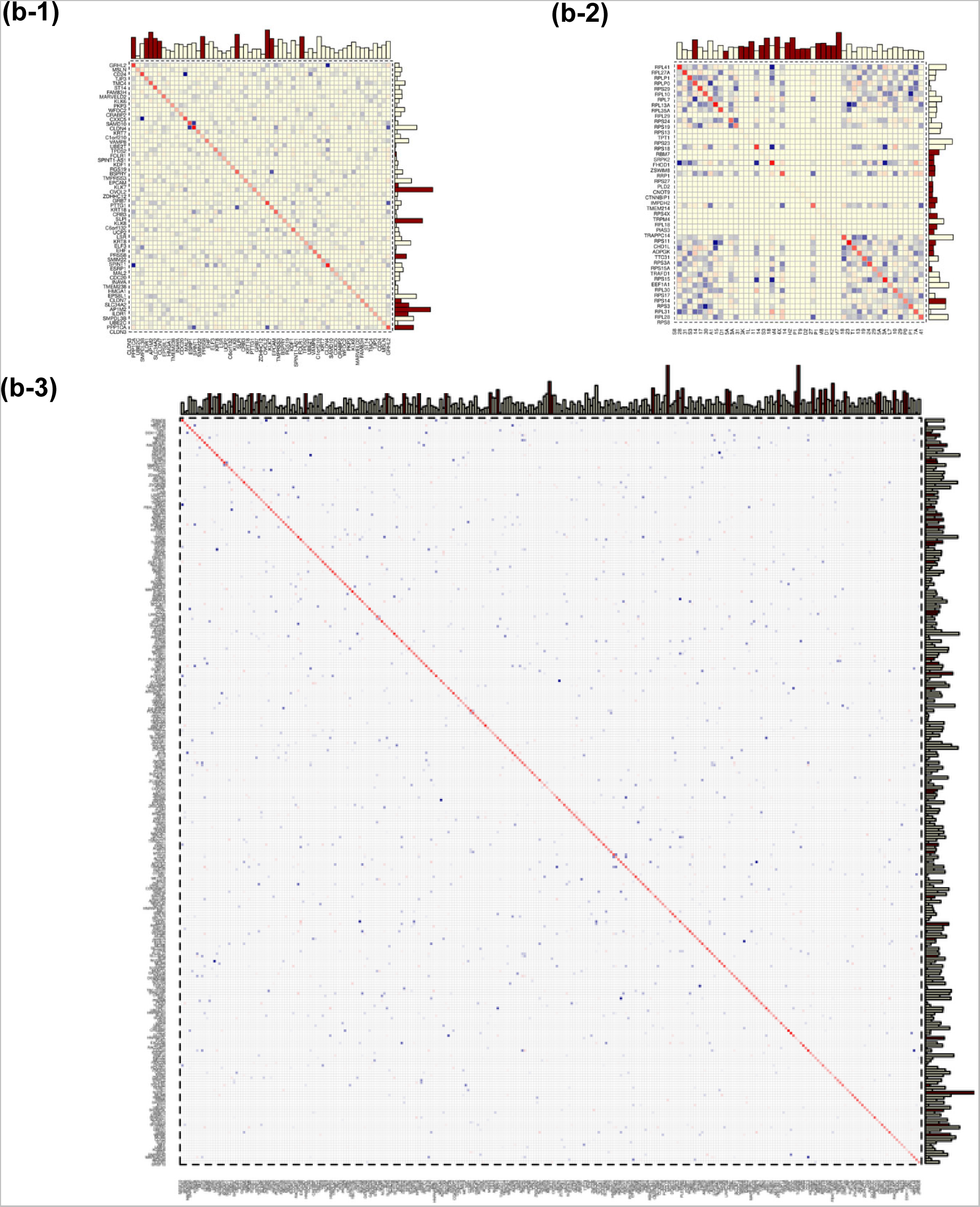
**part I.** Network topologies of the selected PGNs and strong/weak genes therein. Red nodes represent strong genes and yellow nodes represent weak genes. Node sizes are proportional to their marginal effect sizes. Solid line edges represent positive regulations, dotted line edges represent negative regulations. Strength of connections is represented by the width of the edges. **(a-1)** Cancer occurrence. **(a-2)** 5-year survival. **(a-3)** Clinical stage. Figure 4, **part II.** Precision matrix plots of the selected strong and weak genes. Each off diagonal cell in a matrix plot indicates the partial correlation (or coregulation) between the two genes indexed by the corresponding row and column names. Red indicates a positive regulation and blue indicates a negative regulation. The strength of the regulation is indicated by the color scale. The top horizontal histogram depicts the marginal differentiating effect of each gene. The right vertical histogram depicts the network-adjusted differentiating effect (PS) of each gene. Red bars indicate strong genes. Yellow bars indicate regulating weak genes.

We confirmed strong genes previously reported. *SMPDL3B* (marginal p-value=8.8×10^-182^, network-adjusted permutation p-value < 10^-4^) and *SLC34A2* (marginal p-value=1.9×10^-190^, network-adjusted permutation p-value < 10^-4^) were confirmed in the OC occurrence study, *TRAFD1* (marginal p-value = 0.0013, network-adjusted permutation p-value < 10^-4^) and *CHD1L* (marginal p-value = 0.0021, network-adjusted permutation p-value < 10^-4^) were confirmed in the 5-year survival study, and *FANCI* (marginal p-value = 0.0016, network-adjusted permutation p-value < 10^-4^) was confirmed in the clinical stage study. Expression of *SMPDL3B* was found related to specific aptamers for ovarian tumors, such as AptaC2 and AptaC4, through molecular docking [34]. *SLC34A2* overexpression was reported related to development and progression of OC, brain cancer, and pancreatic cancer [35]. *TRAFD1* suppression was observed in ovarian, colon, brain, and renal cancers [36]. *CHD1L* overexpression was reported to augment ovarian carcinoma metastasis [15]. *FANCI* has recently been identified as a new ovarian cancer predisposing gene [37,38]. We also discovered some strong genes not been reported before to be associated with OC. For example, *ILDR1* (marginal p-value = 2.1×10^-204^, network-adjusted permutation p-value < 10^-4^) in OC occurrence study, and *TRAPPC14* (marginal p-value = 5.6×10^-4^, network-adjusted permutation p-value = 2×10^-4^), *RRP1* (marginal p-value = 0.0019, network-adjusted permutation p-value = 6×10^-4^), and *ZSWIM8* (marginal p-value = 0.0045, network-adjusted permutation p-value = 6×10^-4^) in 5-year survival study.

We also identified weak genes in regulations with the strong genes, such as *PPP1CA* (marginal p-value = 3.3×10^-9^, network-adjusted permutation p-value < 10^-4^) and *HMGA1* (marginal p-value = 1.0×10^-57^, network-adjusted permutation p-value < 10^-4^) in occurrence study, *RPS8* (marginal p-value = 0.044, network-adjusted permutation p-value = 0.016), *RPL28* (marginal p-value = 0.36, network-adjusted permutation p-value = 0.024), and *RPL31* (marginal p-value = 0.042, network-adjusted permutation p-value = 0.18) in 5-year survival study, and *MAPKAPK5* (marginal p-value = 0.0017, network-adjusted permutation p-value < 10^-4^) and *BYSL* (marginal p-value = 0.042, network-adjusted permutation p-value < 10^-4^) in the clinical stage study. *PPP1CA* is a catalytic subunit gene and plays an essential role in the growth of cancer cells [39]. *HMGA1* plays a crucial role in the self-renewal and drug resistance of ovarian cancer stem cells [40]. Ribosomal genes, including *RPS8*, *RPL28*, and *RPL31,* have been recently identified as a novel therapeutic target against high-grade OC [41]. Long noncoding RNA *MAPKAPK5- AS1* promotes cancer cell proliferation by cis-regulating the nearby gene MK5 [42]. *BYSL* expression was reported to be elevated and promote tumor cell growth [43].

Several novel OC-associated weak genes that have not been reported in the literature before were identified in our study, such as *GIGYF2 (*marginal p-value = 0.35, network-adjusted permutation p-value < 10^-4^), *GNPAT* (marginal p-value = 0.21, network-adjusted permutation p-value < 10^-4^) and *RAD54L* (marginal p-value = 0.066, network-adjusted permutation p-value < 10^-4^) in the clinical stage study.

**Table 2** lists the top detected PGNs from each of the three studies. Many of these PGNs are overlapping with the top enriched KEGG and/or GO pathways (also see GSEA results). Genes in a KEGG/GO pathway are biologically validated to be related to systematic biology or oncology. Links between genes in a KEGG/GO pathway are lab-confirmed molecular interaction, reaction, and regulations. Overlapping between our selected PGNs and KEGG/GO pathways can serve as biological evidence of our findings. In the OC occurrence study, one of the two netLDA-detected gene networks contains overlapping genes *CLDN7* (weak), *CLDN4* (weak), *CLDN3* (strong) that are also in the *tight junction* pathway (enrichment p-value=1.86×10^- 3^)*, leukocyte transendothelial migration* pathway (enrichment p-value=1.76×10^-3^)*, and cell adhesion molecules cams* pathway (enrichment p-value=2.68×10^-3^). Weak genes *TJP3* and *CRB3,* in the same network, are also overlapped in the *tight junction* pathway. The other predictive gene network selected in OC occurrence study contains two weak genes *PTTG1* and *CDC20* that are overlapping with *cell cycle* pathway (enrichment p-value=1.85×10^-5^). In the survival study, one detected predictive gene network is largely overlapped with *ribosome* pathway (enrichment p-value=1.97×10^-30^). Twenty-eight out of thirty-one genes in the network are in the *ribosome* pathway, which accounts for 31.8% of the 88 leading genes in the ribosome pathway). In the clinical stage study, multiple detected gene networks overlap with *KEGG cell cycle* pathway (enrichment p-value=4.40×10^-4^), *KEGG pathways in cancer* (enrichment p-value=8.12×10^-3^), *GO DNA replication pathway* (enrichment p-value=6.91×10^-9^), *GO DNA recombination pathway* (enrichment p-value=2.09×10^-8^), and *GO chromosome segregation pathway* (enrichment p-value=1.49×10^-7^). Genes overlapping with *KEGG cell cycle* pathway include *MCM2* (strong), *CREBBP* (weak), *ABL1* (weak), *PLK1* (weak), *MCM4* (weak), *MCM6* (weak), *BUB1* (weak), *CDC20* (weak), *CCNB1* (weak), *ORC3* (weak), *SMAD2* (weak), *GSK3B* (weak), and *PCNA* (weak). Genes overlapping with *KEGG pathways in cancer* include *PTGS2* (strong), *KRAS* (strong), *WNT6* (strong), *ABL1* (weak)*, MTOR* (weak)*, SMAD2* (weak)*, STK4* (weak)*, CREBBP* (weak)*, MSH3* (weak)*, RXRB* (weak), and *GSK3B* (weak). Genes overlapping with *GO DNA replication pathway* include *PRIM1* (strong), *MCM6* (weak), *DDX23* (weak), and *WDHD1* (weak). Genes overlapping with *GO DNA recombination pathway* include *MCM2* (strong), *MCM4* (weak), *HMCES* (weak), and *RUVBL1* (weak). Genes overlapping with *GO chromosome segregation pathway* include *BUB1* (weak), *PRC1* (weak), *KIF2C* (weak), *CDC20* (weak), *PLK1* (weak), and *RMI2* (weak).

### Prediction performance comparison with competing ML methods

ROC curves in each study are depicted in **Figure 5**. In the occurrence study, all methods gave almost perfect prediction results – area under the ROC curve (AUC) equaling 1 – as all top genes (strong and weak) have much significant differentiating effects compared to top genes in the survival and clinical stage studies. In the survival study, netLDA gave an AUC = 0.91, much higher than using only the strong genes and Lasso/Ridge/elasticNet (0.85-0.87). XGboosting gave a comparable AUC of 0.90. In the clinical stage study, netLDA also gave the highest AUC = 0.65, XGboosting gave an AUC = 0.61, and Lasso/Ridge/elasticNet and LDA using only the strong genes gave an AUC around 0.5, similar to a random guess.

**Figure 5.**
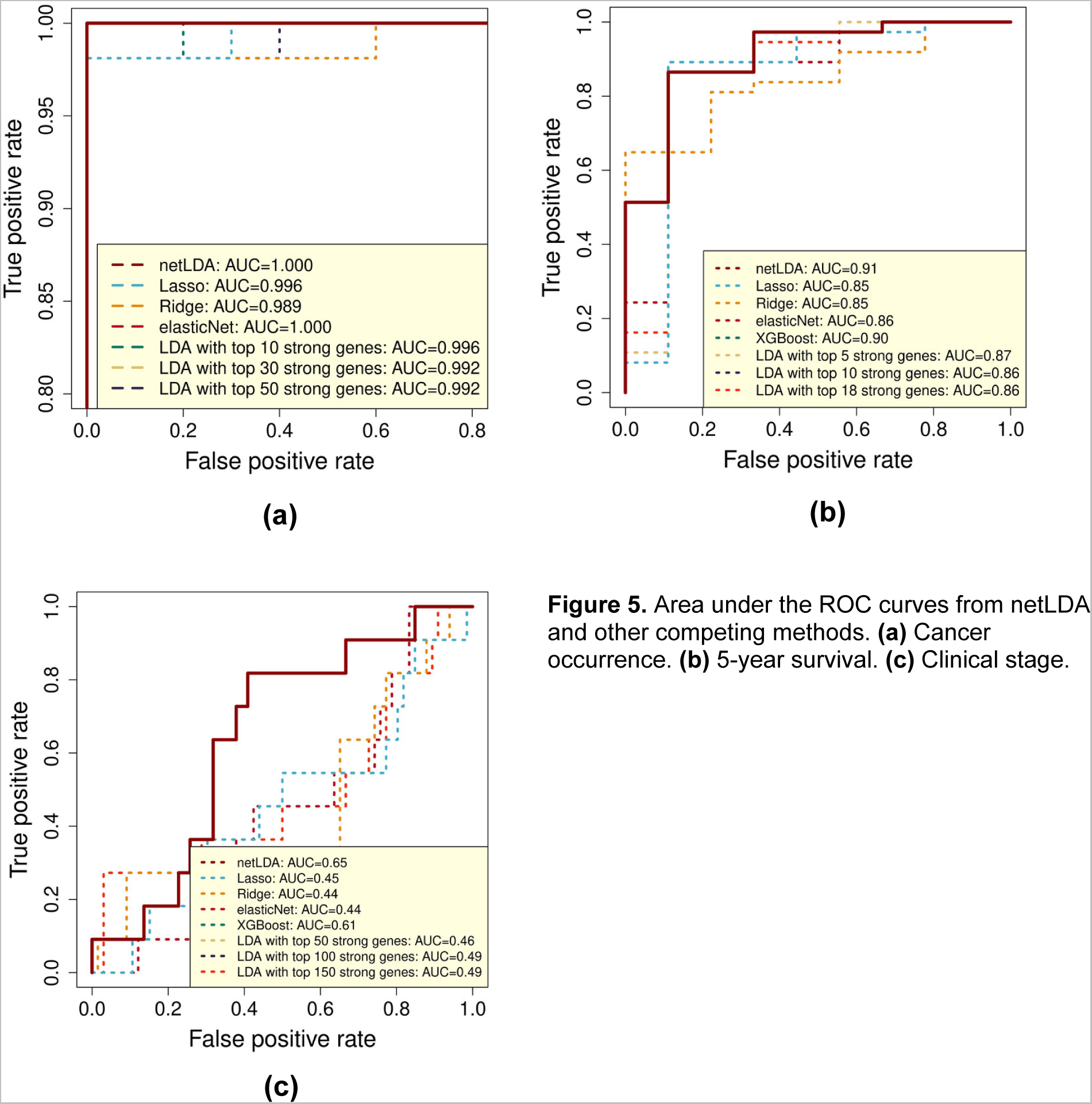
Area under the ROC curves from netLDA and other competing methods. **(a)** Cancer occurrence. **(b)** 5-year survival. **(c)** Clinical stage.

### KM analysis

Figure 6 shows the Kepler-Meijer curves and log-rank test results in the 5-year survival study. Figure 6 **(a)** is for the KM curves and log-rank test between the two netLDA predicted groups using both selected strong/weak genes, and PGN structures. Figure 6 **(b-d)** are KM curves and log-rank tests between high- and low-expression (above and below the median expression value) groups of top three selected strong genes. The two KM curves were more separated, and the log-rank test p-value were more significant between the netLDA predicted groups than those between the expression level groups from a single strong gene, demonstrating the effects of weak genes and PGNs] in improving the classification results.

**Figure 6.**
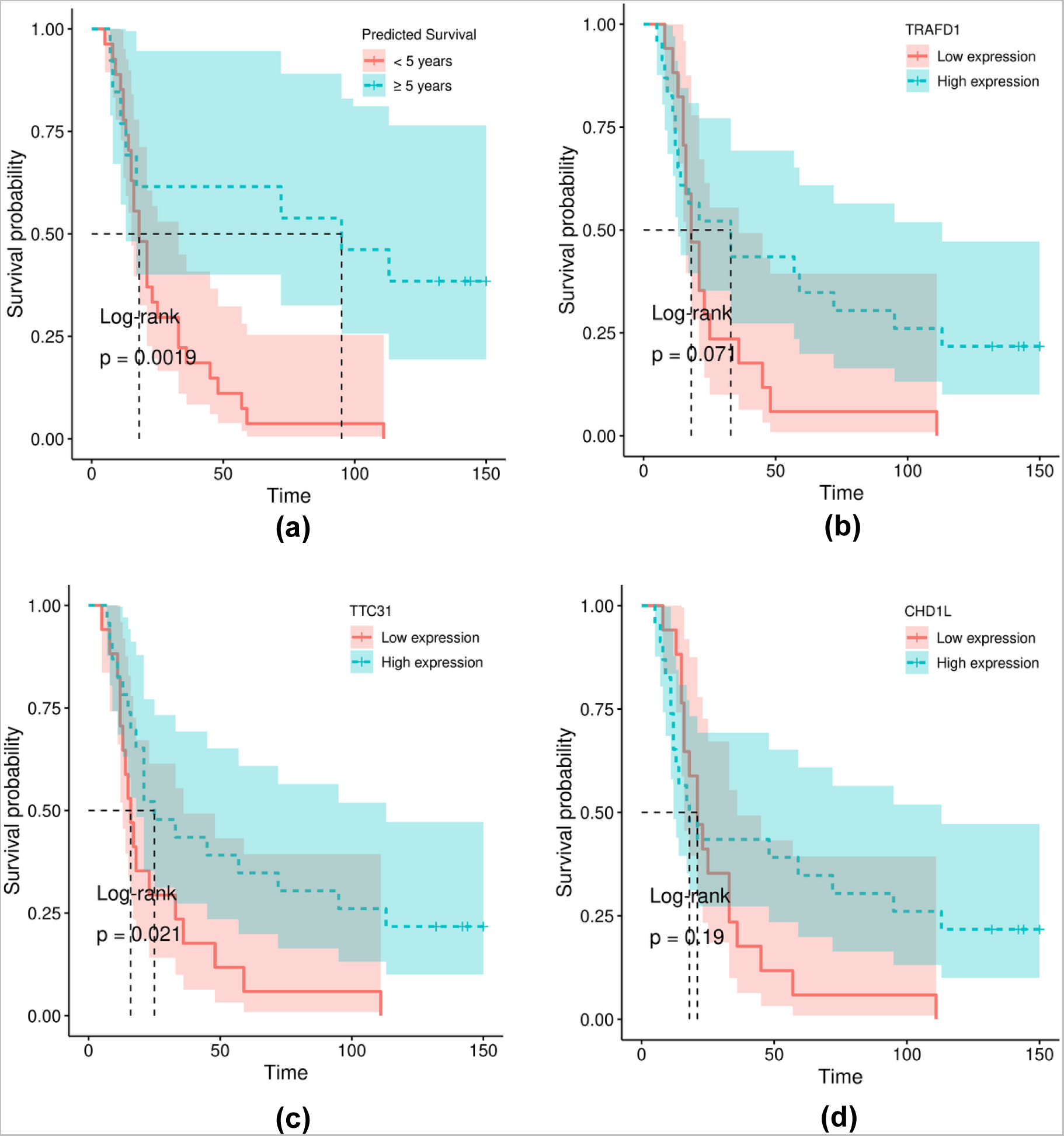
Kepler-Meijer curves and log-rank test results. **(a)** KM curves and log-rank test between the two netLDA predicted groups: long survival (≥ 5 years) and short survival (< 5 years). **(b-d)** KM curves and log-rank tests between high- and low-expression level (above and below the median expression value) of top selected strong genes. **(b)** For gene *TRAFD1*. **(c)** For gene *TTC31*. **(d)** For gene *CHD1L*.

### GSEA

Figure 7 shows KEGG and GO pathway enrichment analysis results. The left panels are examples of the top enriched KEGG pathways for each of the three studies. Tight junction pathway was enriched in the OC occurrence study (enrichment p-value = 1.86×10^-3^), ribosome pathway was enriched in the 5-year survival study (enrichment p-value = 1.97×10^-30^), cell cycle pathway was enriched in the clinical stage study (enrichment p-value = 4.40×10^-4^). Many of the weak genes (highlighted in yellow) overlap with these top enriched pathways, confirming that the weak genes play a biological role in the development and progression of OC. A complete list of enriched KEGG pathways is given in the appendix. The right panels in Figure 7 list the top enriched GO pathways for each study. Top GO pathways enriched in the occurrence study include cell-cell junction organization (enrichment p-value = 4.26×10^-10^), tight junction assembly pathway (enrichment p-value = 3.55×10^-9^), and epidermis development pathway (enrichment p-value = 7.00×10^-9^). Top GO pathways enriched in the survival study include SRP-dependent cotranslational protein targeting to membrane (enrichment p-value = 2.54×10^-55^) and nuclear-transcribed mRNA catabolic process (enrichment p-value = 2.16×10^-53^). Top GO pathways enriched in the clinical stage study include DNA replication (enrichment p-value = 6.91×10^-9^) and recombinational repair (enrichment p-value = 1.59×10^-7^). **Table 3** lists the top enriched KEGG pathways. Lists of top enriched GO pathways are provided in the Supplemental Materials.

**Figure 7.**
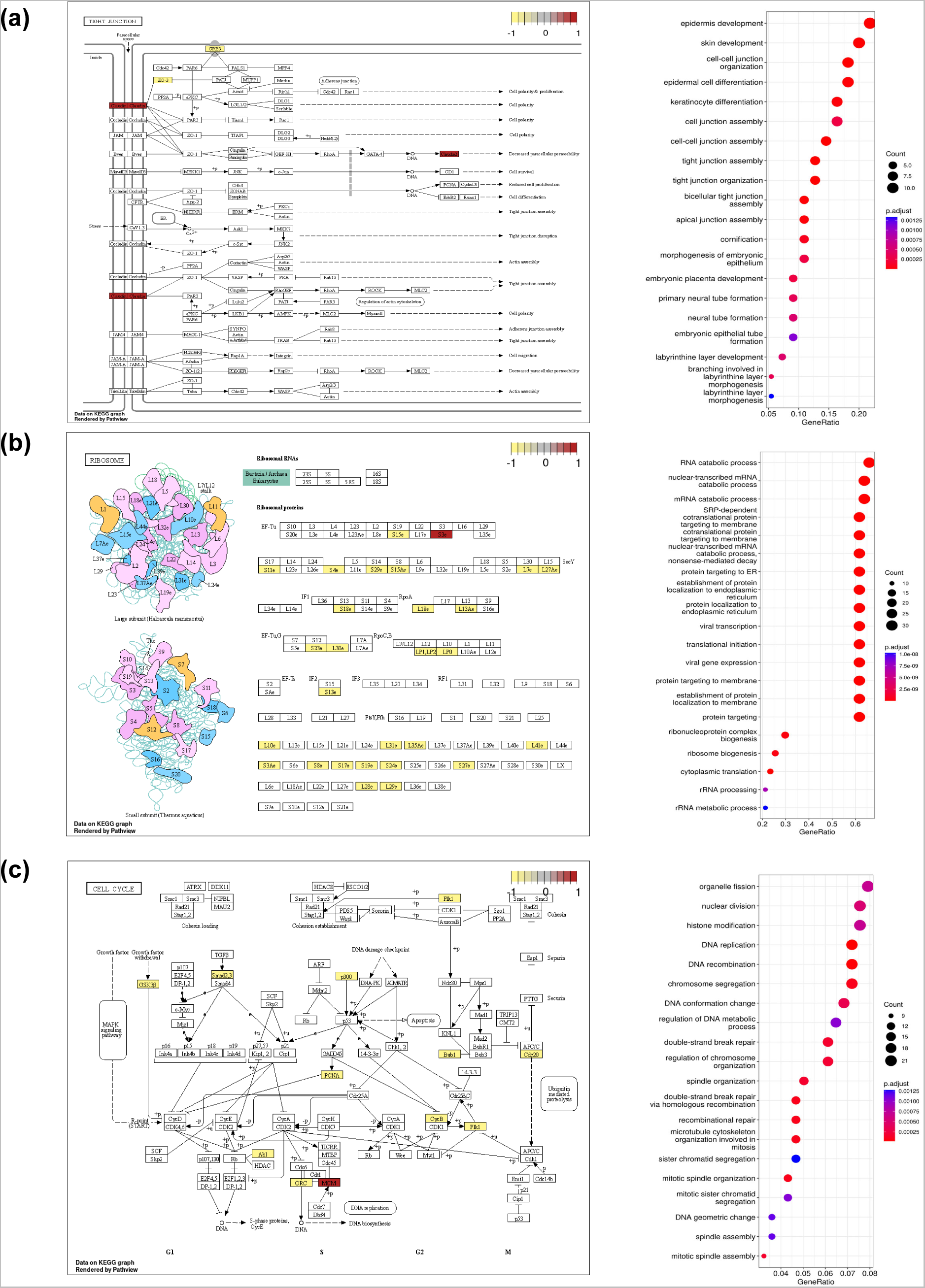
Gene set enrichment analysis results. Left panels: Example top enriched KEGG pathways and genes therein selected by netLDA. Red: selected as strong genes. Yellow: selected as weak genes. **(a)** Tight Junction pathway (p-value=1.9×10^-3^). **(b)** Ribosome pathway (p-value=2.0×10^-30^). **(c)** Cell Cycle pathway (p-value=4.4×10^-4^). Right panels: Top enriched GO pathways. For both left and right panels, **(a)** Cancer occurrence. **(b)** 5-year survival. **(c)** Clinical stage.

**Table 3.**
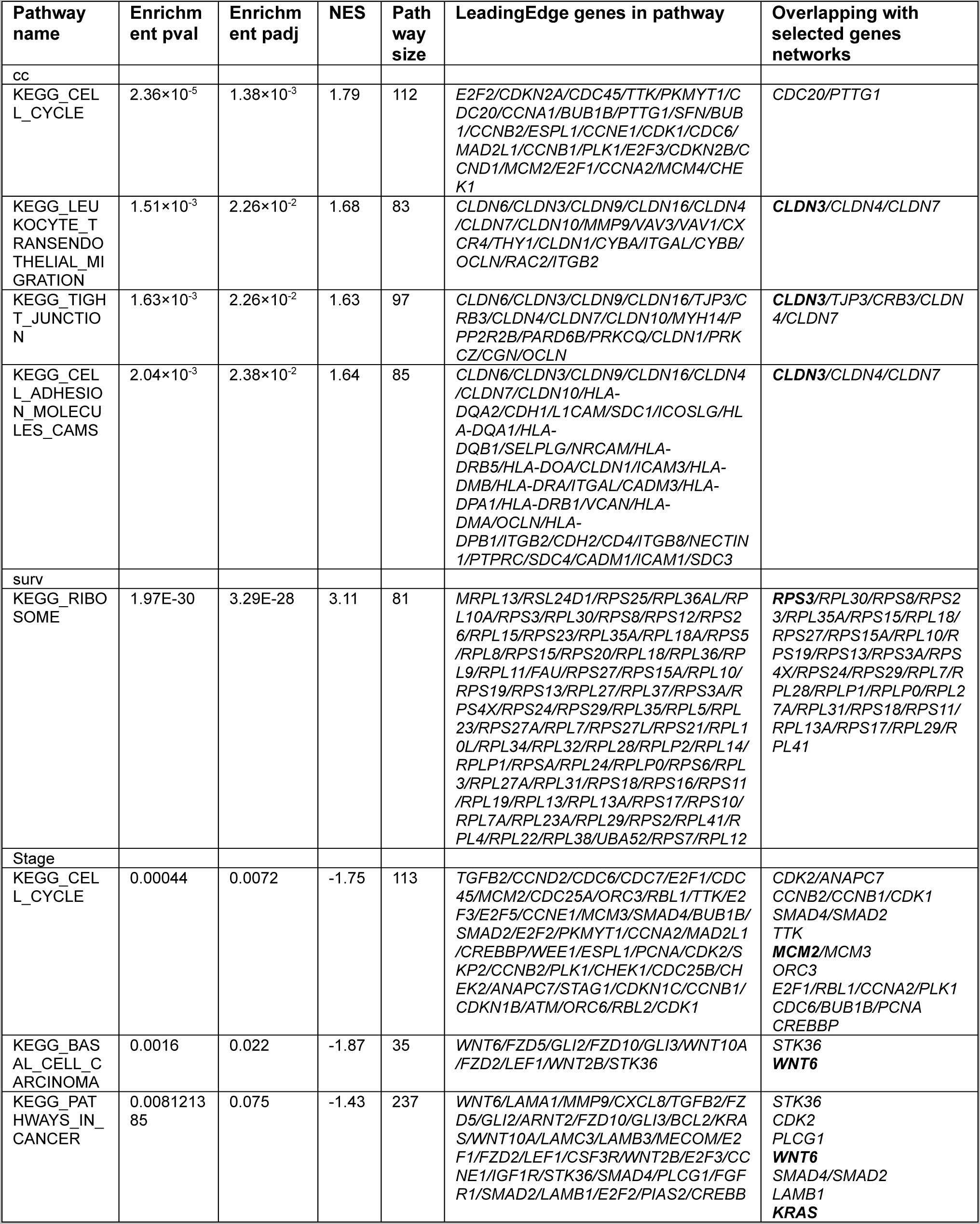
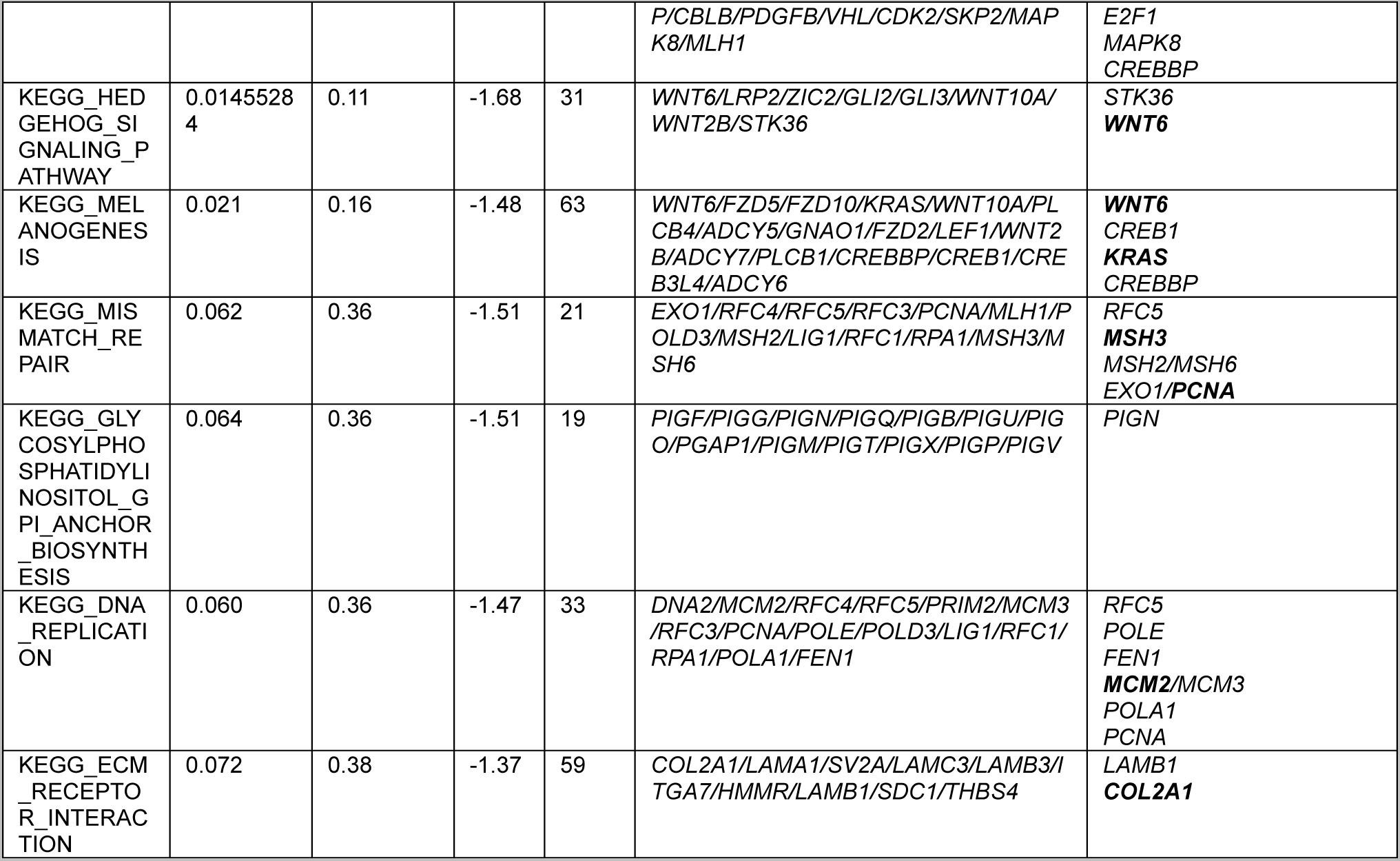
Top enriched KEGG pathways and their overlapping selected gene networks.

### Cellular mapping for the selected genes using scRNAseq data

Figure 8 shows the cellular distribution of the GSE229343 scRNAseq data and the expression maps of the selected feature genes. In Figure 8 **(a)**, Seurat was first used to identify 28 cell subtype clusters using resolution = 0.2. For OC occurrence study, many genes are expressed on epithelial cells (including strong genes: *CLDN3*, *SLC34A2, SMIM22, FOLR1*, and weak genes: *CLDN7, MAL2, SPRINT1, PRSS8, EHF, ELF3, KRT8, SLPI, KRT18, EPCAM, VAMP8, KRT7, CLDN4, WFDC2, CD24, MSLN*); on fore/mid/hindgut epithelial cells (including strong genes: *CLDN3*, *SMIM22,* and weak genes: *CLDN7, MAL2, SPINT1, PRSS8, ELF3, KRT8, SLPI, KRT18, EPCAM, KRT7, CLDN4*); on cycling neural program/mesenchymal stem cells (including weak genes: *UBE2C, CDC20, PTTG1, UBE2T*); on airway/retinal epi/ciliated cells (including strong genes: *CLDN3*, *SMIM22, FOLR1,* and weak genes: *ELF3, KRT8, UCP2, SLPI, EPCAM, KRT7, CLDN4, WFDC2, CD24*); on myeloid/T cells (including weak genes: *UCP2, VAMP8*); and on immature neuron cells (including weak gene CD24). For the clinical stage study, many genes are overexpressed on cycling neural program/Mesenchymal stem cells (including weak genes: *CDC20, UBE2T, NUSAP1, CCNB1, PLK1, ASPM, PRC1, KIF20A*); on immature neuron cells (including strong gene TUBB2B); and on myeloid cells (including weak gene CD163). For the survival study, as there were very few genes mapped to the scRNAseq GSE229343 gene set, we did not observe a particular cellular express pattern for the selected genes.

**Figure 8.**
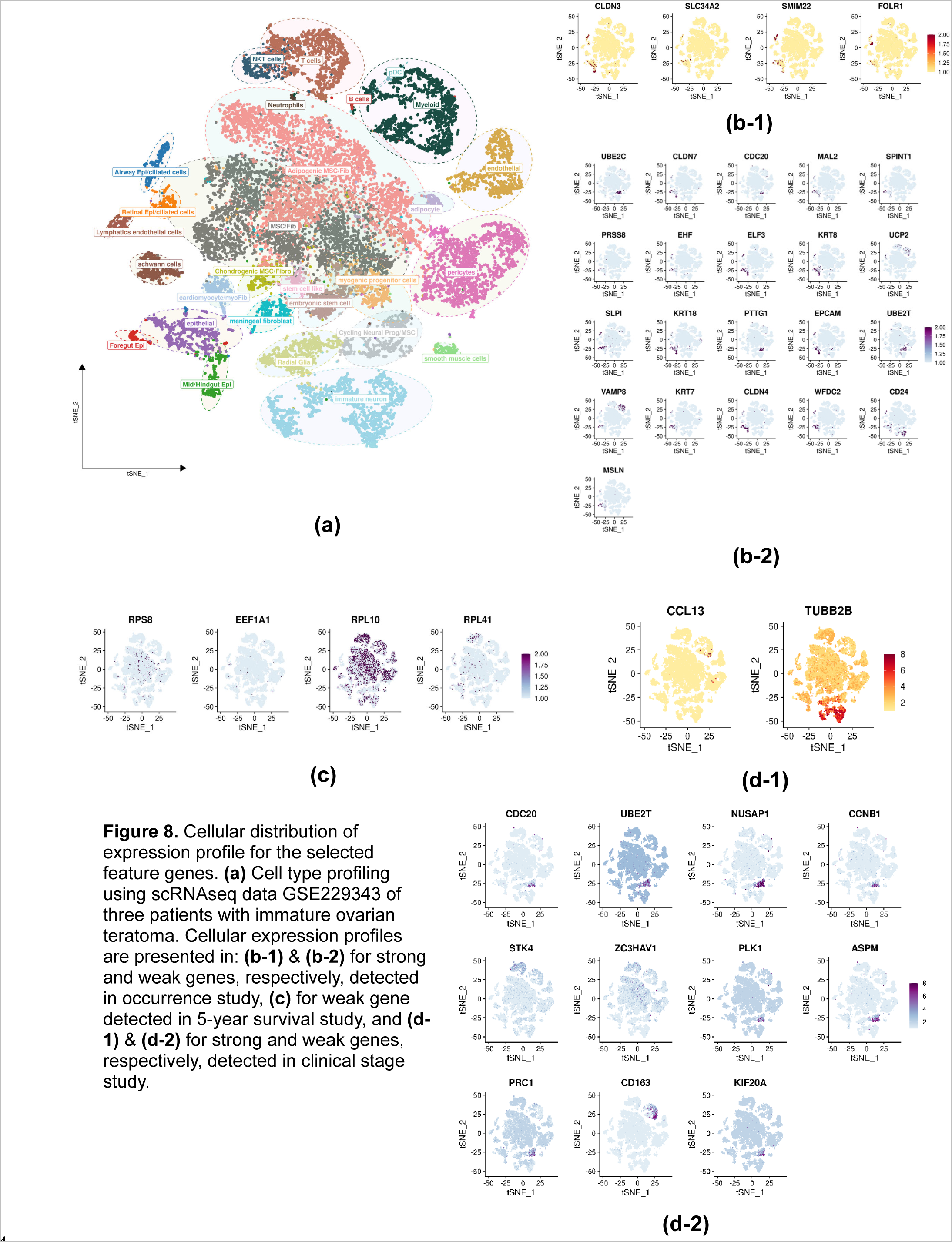
Cellular distribution of expression profile for the selected feature genes. **(a)** Cell type profiling using scRNAseq data GSE229343 of three patients with immature ovarian teratoma. Cellular expression profiles are presented in: **(b-1)** & **(b-2)** for strong and weak genes, respectively, detected in occurrence study, **(c)** for weak gene detected in 5-year survival study, and **(d-1)** & **(d-2)** for strong and weak genes, respectively, detected in clinical stage study.

## Summary and Discussion

We confirmed previously identified prognostic genes such as *EPCAM, UBE2C, CHD1L, TP53*, *CD24* [53], *WFDC2* [53], and *FANCI* associated with OC occurrence, survival, or clinical stage. We identified novel susceptibility strong genes including: *ILDR1* in ocurrence study*, TRAPPC14, RRP1,* and *ZSWIM8* in the survival study, as well as novel coregulating weak genes including *GIGYF2, GNPAT, RAD54L*, and *ELL* in the clinical stage study. Our identified gene networks overlapped with KEGG *tight junction, leukocyte transendothelial migration,* and *cell cycle* pathways in the occurrence study; with *ribosome* pathway in the survival study; and with *cell cycle* pathway and *pathways in cancer* in the clinical stage study. We found many identified genes particularly expressed on epithelial cells in the occurrence study and on cycling neural program/Mesenchymal stem cells in the clinical stage study. By incorporating gene network structures and weak genes, netLDA significantly improves the prediction performance compared to other ML/DL methods such as Lasso, Ridge, elasticNet, XGboost, and LDA with only strong genes.

A major contribution of this work is the identification of prognostic oncogenes, especially weak genes in the OC related pathways. The *CLDN* genes (*CLDN7* and *CLDN4* detected in the OC occurrence study) in the *tight junction* pathway and *cell adhesion molecules cams* pathway, which functioning as one of the protective barriers in the epithelial and endothelial cells, were also observed to overexpress on fore/mid/hindgut epithelial cells in the single cell analysis. The weak genes detected in the survival study, mainly ribosomal genes, such as RPS8 and RPL28, overlapping with *ribosome* pathway, which was known to promote protein homeostasis in cancers by fine-tuning protein synthesis and preventing toxic protein aggregation [102]. The *CCN* gene family (*CCNB1* detected in the clinical stage study) and CDK genes (*CDK2* within the detected network indexed by *PRIM1* in clinical stage study, see **Table 2**, even though it is not in the final selected weak gene set) are coregulating genes in the *cell cycle* pathway. These regulating weak genes were not reported in the literature, as they were difficult to detect by themselves.

Although netLDA incorporates gene coregulation network structures into calculating gene differential expressions, it assumes the same network structures (topology, coregulation direction and strength, etc.) between different outcome groups. In real applications, the network structures can be different between groups, in which case a quadratic discriminant analysis might be a better approach. However, accurate and robust inference of different network structures requires large sample sizes per group. Moreover, gene coregulations are usually more stable and robust compared to individual gene expression levels. That is, even individual gene expression levels can vary a lot between different disease groups, cell types, and environments, but the gene-gene coregulation maintains rather stable across the heterogeneous situations. Such robustness in gene networks is critical for assembling a dynamic biology system. Essentially, the network structures inferred by netLDA are the average of different groups when the same network structure assumption is violated.

Most of the weak genes selected in the OC occurrence study also have strong marginal DE effects. In that sense, they may also be considered as strong genes. The prediction performance is dominated by the top strong genes; therefore, the prediction performance (Figure 5) was not much different between netLDA and LDA using only strong genes and other competing methods. The predictive effects of weak genes were manifested in the survival and clinical stage studies, where the marginal DE differential effects of genes are much weaker than that in the occurrence study (Figure 3 and Figure 5). Especially in the clinical stage study, as the strong genes only explained a small portion of the outcome variation on the training data, netLDA dug deeper with more weak genes residing in more predictive networks compared to the occurrence and survival study, in order to accumulate sufficient information to optimize the prediction accuracy on the test data.

The selected gene sets from the occurrence, survival, and stage studies are non-overlapping. This is mainly because the input gene sets are different for the three studies. Moreover, it also reveals that the OC development and progression may have different underlying molecular mechanisms.

Since gene expressions in bulk RNAseq data are averaged expressions over different types of cells, DE effects of some genes might be washed out in the averaging. For example, a gene that is significantly differentially expressed only on a particular cell type but not on other cell types might exhibit only a marginally weak DE effect. A gene differentially expressed on two cell types but in different directions will show no marginal DE effect due to signal cancellation. The netLDA is a desired method for detecting such genes. It would be an optimal validation approach to confirm the DE effects of the selected genes using scRNAseq data from both cohorts. Our feature mapping of the selected genes using scRNAseq data helped identify which types of cells the genes are particularly expressed on. Investigators are recommended to conduct DE analyses using scRNAseq data on the identified cell types to further validate the findings.

## Data Availability

All data produced are available online at:
The Cancer Genome Atlas (TCGA) (https://xenabrowser.net/);
The Genotype-Tissue Expression (GTEx) (https://www.gtexportal.org/);
Gene Expression Omnibus (GEO) (https://www.ncbi.nlm.nih.gov/geo/)

## Abbreviations

DE: Differentially expressed
GEO: Gene Expression Omnibus
GO: GeneOntology
GSEA: Gene set enrichment analysis
GTEx: The Genotype-Tissue Expression project
KEGG: Kyoto Encyclopedia of Genes and Genomes
KM: Kepler-Meijer
LDA: Linear discriminant analysis
OC: Ovarian cancer
PGN: Predictive gene networks
RNAseq: RNA sequencing
scRNAseq: single cell RNA sequencing
TCGA: The Cancer Genome Atlas

## Reference

1. Matulonis UA, Sood AK, Fallowfield L, Howitt BE, Sehouli J, Karlan BY. Ovarian cancer. Nat Rev Dis Primers. 2016 Aug 25;2:16061. doi: 10.1038/nrdp.2016.61. PMID: 27558151; PMCID: PMC7290868.

2. Stewart C, Ralyea C, Lockwood S. Ovarian Cancer: An Integrated Review. Semin Oncol Nurs. 2019 Apr;35(2):151–156. doi: 10.1016/j.soncn.2019.02.001. Epub 2019 Mar 11. PMID: 30867104.

3. Alexandrova E, Pecoraro G, Sellitto A, Melone V, Ferravante C, Rocco T, Guacci A, Giurato G, Nassa G, Rizzo F, Weisz A, Tarallo R. An Overview of Candidate Therapeutic Target Genes in Ovarian Cancer. Cancers (Basel). 2020 Jun 4;12(6):1470. doi: 10.3390/cancers12061470. PMID: 32512900; PMCID: PMC7352306.

4. National Cancer Institute. Surveillance, Epidemiology, and End Results Program. https://seer.cancer.gov/statfacts/html/ovary.html.

5. Petrucelli N, Daly MB, Pal T. BRCA1- and BRCA2-Associated Hereditary Breast and Ovarian Cancer. 1998 Sep 4 [updated 2023 Sep 21]. In: Adam MP, Mirzaa GM, Pagon RA, Wallace SE, Bean LJH, Gripp KW, Amemiya A, editors. GeneReviews® [Internet]. Seattle (WA): University of Washington, Seattle; 1993–2023. PMID: 20301425.

6. Sánchez-Lorenzo L, Salas-Benito D, Villamayor J, Patiño-García A, González-Martín A. The BRCA Gene in Epithelial Ovarian Cancer. Cancers (Basel). 2022 Feb 27;14(5):1235. doi: 10.3390/cancers14051235. PMID: 35267543; PMCID: PMC8909050.

7. Casaubon JT, Kashyap S, Regan JP. BRCA1 and BRCA2 Mutations. 2023 Jul 23. In: StatPearls [Internet]. Treasure Island (FL): StatPearls Publishing; 2023 Jan–. PMID: 29262038.

8. Takaoka M, Miki Y. BRCA1 gene: function and deficiency. Int J Clin Oncol. 2018 Feb;23(1):36–44. doi: 10.1007/s10147-017-1182-2. Epub 2017 Sep 7. PMID: 28884397.

9. Tayama S, Motohara T, Narantuya D, Li C, Fujimoto K, Sakaguchi I, Tashiro H, Saya H, Nagano O, Katabuchi H. The impact of EpCAM expression on response to chemotherapy and clinical outcomes in patients with epithelial ovarian cancer. Oncotarget. 2017 Jul 4;8(27):44312–44325. doi: 10.18632/oncotarget.17871. PMID: 28574829; PMCID: PMC5546482.

10. Nunna S, Reinhardt R, Ragozin S, Jeltsch A. Targeted methylation of the epithelial cell adhesion molecule (EpCAM) promoter to silence its expression in ovarian cancer cells. PLoS One. 2014 Jan 29;9(1):e87703. doi: 10.1371/journal.pone.0087703. PMID: 24489952; PMCID: PMC3906225.

11. Zhang Y, Cao L, Nguyen D, Lu H. TP53 mutations in epithelial ovarian cancer. Transl Cancer Res. 2016 Dec;5(6):650–663. doi: 10.21037/tcr.2016.08.40. PMID: 30613473; PMCID: PMC6320227.

12. Silwal-Pandit L, Langerød A, Børresen-Dale AL. TP53 Mutations in Breast and Ovarian Cancer. Cold Spring Harb Perspect Med. 2017 Jan 3;7(1):a026252. doi: 10.1101/cshperspect.a026252. PMID: 27815305; PMCID: PMC5204332.

13. He WP, Guo YY, Yang GP, Lai HL, Sun TT, Zhang ZW, Ouyang LL, Zheng Y, Tian LM, Li XH, You ZS, Xie D, Yang GF. CHD1L promotes EOC cell invasiveness and metastasis via the regulation of METAP2. Int J Med Sci. 2020 Aug 29;17(15):2387–2395. doi: 10.7150/ijms.48615. PMID: 32922205; PMCID: PMC7484650.

14. Soltan MA, Eldeen MA, Eid RA, Alyamani NM, Alqahtani LS, Albogami S, Jafri I, Park MN, Alsharif G, Fayad E, Mohamed G, Osman R, Kim B, Zaki MSA. A pan-cancer analysis reveals CHD1L as a prognostic and immunological biomarker in several human cancers. Front Mol Biosci. 2023 Mar 23;10:1017148. doi: 10.3389/fmolb.2023.1017148. PMID: 37033447; PMCID: PMC10076660.

15. He WP, Zhou J, Cai MY, Xiao XS, Liao YJ, Kung HF, Guan XY, Xie D, Yang GF. CHD1L protein is overexpressed in human ovarian carcinomas and is a novel predictive biomarker for patients survival. BMC Cancer. 2012 Sep 29;12:437. doi: 10.1186/1471-2407-12-437. PMID: 23020525; PMCID: PMC3551745.

16. Pawar A, Chowdhury OR, Chauhan R, Talole S, Bhattacharjee A. Identification of key gene signatures for the overall survival of ovarian cancer. J Ovarian Res. 2022 Jan 20;15(1):12. doi: 10.1186/s13048-022-00942-0. PMID: 35057823; PMCID: PMC8780391.

17. Millstein J, Budden T, Goode EL, Anglesio MS, Talhouk A, Intermaggio MP, Leong HS, Chen S, Elatre W, Gilks B, Nazeran T, Volchek M, Bentley RC, Wang C, Chiu DS, Kommoss S, Leung SCY, Senz J, Lum A, Chow V, Sudderuddin H, Mackenzie R, George J; AOCS Group; Fereday S, Hendley J, Traficante N, Steed H, Koziak JM, Köbel M, McNeish IA, Goranova T, Ennis D, Macintyre G, Silva De Silva D, Ramón Y Cajal T, García-Donas J, Hernando Polo S, Rodriguez GC, Cushing-Haugen KL, Harris HR, Greene CS, Zelaya RA, Behrens S, Fortner RT, Sinn P, Herpel E, Lester J, Lubiński J, Oszurek O, Tołoczko A, Cybulski C, Menkiszak J, Pearce CL, Pike MC, Tseng C, Alsop J, Rhenius V, Song H, Jimenez-Linan M, Piskorz AM, Gentry-Maharaj A, Karpinskyj C, Widschwendter M, Singh N, Kennedy CJ, Sharma R, Harnett PR, Gao B, Johnatty SE, Sayer R, Boros J, Winham SJ, Keeney GL, Kaufmann SH, Larson MC, Luk H, Hernandez BY, Thompson PJ, Wilkens LR, Carney ME, Trabert B, Lissowska J, Brinton L, Sherman ME, Bodelon C, Hinsley S, Lewsley LA, Glasspool R, Banerjee SN, Stronach EA, Haluska P, Ray-Coquard I, Mahner S, Winterhoff B, Slamon D, Levine DA, Kelemen LE, Benitez J, Chang-Claude J, Gronwald J, Wu AH, Menon U, Goodman MT, Schildkraut JM, Wentzensen N, Brown R, Berchuck A, Chenevix-Trench G, deFazio A, Gayther SA, García MJ, Henderson MJ, Rossing MA, Beeghly-Fadiel A, Fasching PA, Orsulic S, Karlan BY, Konecny GE, Huntsman DG, Bowtell DD, Brenton JD, Doherty JA, Pharoah PDP, Ramus SJ. Prognostic gene expression signature for high-grade serous ovarian cancer. Ann Oncol. 2020 Sep;31(9):1240–1250. doi: 10.1016/j.annonc.2020.05.019. Epub 2020 May 28. PMID: 32473302; PMCID: PMC7484370.

18. Marchini S, Mariani P, Chiorino G, Marrazzo E, Bonomi R, Fruscio R, Clivio L, Garbi A, Torri V, Cinquini M, Dell’Anna T, Apolone G, Broggini M, D’Incalci M. Analysis of gene expression in early-stage ovarian cancer. Clin Cancer Res. 2008 Dec 1;14(23):7850–60. doi: 10.1158/1078-0432.CCR-08-0523. PMID: 19047114.

19. Li Y, Hong HG, Ahmed SE, Li Y. Weak signals in high-dimension regression: detection, estimation and prediction. Appl Stoch Models Bus Ind. 2019;35(2):283–98. Epub 2019/11/02. doi: 10.1002/asmb.2340. PubMed PMID: 31666801; PubMed Central PMCID: PMCPMC6821396.

20. Li Y, Hong HG, Li Y. Multiclass linear discriminant analysis with ultrahigh-dimensional features. Biometrics. 2019;75(4):1086–97. Epub 2019/04/23. doi: 10.1111/biom.13065. PubMed PMID: 31009070; PubMed Central PMCID: PMCPMC6810714.

21. Li Y. (2020). A Local-Network Guided Linear Discriminant Analysis for Classifying Lung Cancer Subtypes using Individual Genome-Wide Methylation Profiles. In: Arai, K., Bhatia, R., Kapoor, S. (eds) Proceedings of the Future Technologies Conference (FTC) 2019. FTC 2019. Advances in Intelligent Systems and Computing, vol 1069. Springer, Cham. 10.1007/978-3-030-32520-6_50.

22. Li Y, Kang J, Wu C, et al. A machine-learning approach for detection of local brain networks and marginally weak signals identifies novel AD/MCI differentiating connectomic neuroimaging biomarkers. BioRXiv. doi: 10.1101/2021.07.29.454368.

23. Zhou X, Zhang J, Ding Y, Huang H, Li Y, Chen W. Predicting late-stage age-related macular degeneration by integrating marginally weak SNPs in GWA studies. Front Genet. 2023;14:1075824. Epub 2023/04/18. doi: 10.3389/fgene.2023.1075824. PubMed PMID: 37065470; PubMed Central PMCID: PMCPMC10101437.

24. Robinson MD, McCarthy DJ, Smyth GK. edgeR: a Bioconductor package for differential expression analysis of digital gene expression data. Bioinformatics. 2010 Jan 1;26(1):139–40. doi: 10.1093/bioinformatics/btp616. Epub 2009 Nov 11. PMID: 19910308; PMCID: PMC2796818.

25. Satija R, Farrell JA, Gennert D, Schier AF, Regev A. Spatial reconstruction of single-cell gene expression data. Nat Biotechnol. 2015 May;33(5):495–502. doi: 10.1038/nbt.3192. Epub 2015 Apr 13. PMID: 25867923; PMCID: PMC4430369.

26. Tibshirani R. Regression Shrinkage and Selection via the Lasso. Journal of the Royal Statistical Society: Series B (Methodological). 1996; 58, 267–288. http://www.jstor.org/stable/2346178. 10.1111/j.2517-6161.1996.tb02080.x.

27. Hilt DE, Seegrist, DW. Ridge, a computer program for calculating ridge regression estimates. 1977; doi:10.5962/bhl.title.68934.

28. Zou H, Hastie T. Regularization and Variable Selection via the Elastic Net. Journal of the Royal Statistical Society, Series B. 2005; 67 (2): 301–320. CiteSeerX 10.1.1.124.4696. doi:10.1111/j.1467-9868.2005.00503.x. S2CID 122419596.

29. Chen TQ, Guestrin C. XGBoost: A Scalable Tree Boosting System. In Krishnapuram B, Shah M, Smola AJ, Aggarwal CC, Shen D, Rastogi R (eds.). Proceedings of the 22nd ACM SIGKDD International Conference on Knowledge Discovery and Data Mining, San Francisco, CA, USA, 2016 August 13-17, 2016. ACM. pp. 785–794. arXiv:1603.02754. doi:10.1145/2939672.2939785. ISBN 9781450342322. S2CID 4650265.

30. Liu GM, Zeng HD, Zhang CY, Xu JW. Identification of a six-gene signature predicting overall survival for hepatocellular carcinoma. Cancer Cell Int. 2019 May 21;19:138. doi: 10.1186/s12935-019-0858-2. PMID: 31139015; PMCID: PMC6528264.

31. Subramanian A, Tamayo P, Mootha VK, Mukherjee S, Ebert BL, Gillette MA, Paulovich A, Pomeroy SL, Golub TR, Lander ES, Mesirov JP. Gene set enrichment analysis: a knowledge-based approach for interpreting genome-wide expression profiles. Proc Natl Acad Sci U S A. 2005 Oct 25;102(43):15545–50. doi: 10.1073/pnas.0506580102. Epub 2005 Sep 30. PMID: 16199517; PMCID: PMC1239896.

32. Harris MA, Clark J, Ireland A, Lomax J, Ashburner M, Foulger R, Eilbeck K, Lewis S, Marshall B, Mungall C, Richter J, Rubin GM, Blake JA, Bult C, Dolan M, Drabkin H, Eppig JT, Hill DP, Ni L, Ringwald M, Balakrishnan R, Cherry JM, Christie KR, Costanzo MC, Dwight SS, Engel S, Fisk DG, Hirschman JE, Hong EL, Nash RS, Sethuraman A, Theesfeld CL, Botstein D, Dolinski K, Feierbach B, Berardini T, Mundodi S, Rhee SY, Apweiler R, Barrell D, Camon E, Dimmer E, Lee V, Chisholm R, Gaudet P, Kibbe W, Kishore R, Schwarz EM, Sternberg P, Gwinn M, Hannick L, Wortman J, Berriman M, Wood V, de la Cruz N, Tonellato P, Jaiswal P, Seigfried T, White R; Gene Ontology Consortium. The Gene Ontology (GO) database and informatics resource. Nucleic Acids Res. 2004 Jan 1;32(Database issue):D258–61. doi: 10.1093/nar/gkh036. PMID: 14681407; PMCID: PMC308770.

33. Kanehisa M, Goto S. KEGG: kyoto encyclopedia of genes and genomes. Nucleic Acids Res. 2000 Jan 1;28(1):27–30. doi: 10.1093/nar/28.1.27. PMID: 10592173; PMCID: PMC102409.

34. Abreu RDS, Antunes D, Moreira ADS, Passetti F, Mendonça JB, de Araújo NS, Sassaro TF, Alberto AVP, Carrossini N, Fernandes PV, Costa MA, Guimarães ACR, Degrave WMS, Waghabi MC. Next Generation of Ovarian Cancer Detection Using Aptamers. Int J Mol Sci. 2023 Mar 28;24(7):6315. doi: 10.3390/ijms24076315. PMID: 37047289; PMCID: PMC10094455.

35. Vlasenkova R, Nurgalieva A, Akberova N, Bogdanov M, Kiyamova R. Characterization of SLC34A2 as a Potential Prognostic Marker of Oncological Diseases. Biomolecules. 2021 Dec 14;11(12):1878. doi: 10.3390/biom11121878. PMID: 34944522; PMCID: PMC8699446.

36. Sehovic E, Hadrovic A, Dogan S. Detection and analysis of stable and flexible genes towards a genome signature framework in cancer. Bioinformation. 2019 Nov 10;15(10):772–779. doi: 10.6026/97320630015772. PMID: 31831960; PMCID: PMC6900328.

37. Fierheller CT, Alenezi WM, Serruya C, Revil T, Amuzu S, Bedard K, Subramanian DN, Fewings E, Bruce JP, Prokopec S, Bouchard L, Provencher D, Foulkes WD, El Haffaf Z, Mes-Masson AM, Tischkowitz M, Campbell IG, Pugh TJ, Greenwood CMT, Ragoussis J, Tonin PN. Molecular Genetic Characteristics of FANCI, a Proposed New Ovarian Cancer Predisposing Gene. Genes (Basel). 2023 Jan 20;14(2):277. doi: 10.3390/genes14020277. PMID: 36833203; PMCID: PMC9956348.

38. Fierheller CT, Guitton-Sert L, Alenezi WM, Revil T, Oros KK, Gao Y, Bedard K, Arcand SL, Serruya C, Behl S, Meunier L, Fleury H, Fewings E, Subramanian DN, Nadaf J, Bruce JP, Bell R, Provencher D, Foulkes WD, El Haffaf Z, Mes-Masson AM, Majewski J, Pugh TJ, Tischkowitz M, James PA, Campbell IG, Greenwood CMT, Ragoussis J, Masson JY, Tonin PN. A functionally impaired missense variant identified in French Canadian families implicates FANCI as a candidate ovarian cancer-predisposing gene. Genome Med. 2021 Dec 3;13(1):186. doi: 10.1186/s13073-021-00998-5. PMID: 34861889; PMCID: PMC8642877.

39. Takakura S, Kohno T, Manda R, Okamoto A, Tanaka T, Yokota J. Genetic alterations and expression of the protein phosphatase 1 genes in human cancers. Int J Oncol. 2001 Apr;18(4):817–24. doi: 10.3892/ijo.18.4.817. PMID: 11251179.

40. Kim DK, Seo EJ, Choi EJ, Lee SI, Kwon YW, Jang IH, Kim SC, Kim KH, Suh DS, Seong-Jang K, Lee SC, Kim JH. Crucial role of HMGA1 in the self-renewal and drug resistance of ovarian cancer stem cells. Exp Mol Med. 2016 Aug 26;48(8):e255. doi: 10.1038/emm.2016.73. PMID: 27561949; PMCID: PMC5007643.

41. Yan S, Frank D, Son J, Hannan KM, Hannan RD, Chan KT, Pearson RB, Sanij E. The Potential of Targeting Ribosome Biogenesis in High-Grade Serous Ovarian Cancer. Int J Mol Sci. 2017 Jan 20;18(1):210. doi: 10.3390/ijms18010210. PMID: 28117679; PMCID: PMC5297839.

42. Yang T, Chen WC, Shi PC, Liu MR, Jiang T, Song H, Wang JQ, Fan RZ, Pei DS, Song J. Long noncoding RNA MAPKAPK5-AS1 promotes colorectal cancer progression by cis-regulating the nearby gene MK5 and acting as a let-7f-1-3p sponge. J Exp Clin Cancer Res. 2020 Jul 20;39(1):139. doi: 10.1186/s13046-020-01633-8. Erratum in: J Exp Clin Cancer Res. 2022 Dec 13;41(1):341. PMID: 32690100; PMCID: PMC7370515.

43. Gao S, Sha Z, Zhou J, Wu Y, Song Y, Li C, Liu X, Zhang T, Yu R. BYSL contributes to tumor growth by cooperating with the mTORC2 complex in gliomas. Cancer Biol Med. 2021 Feb 15;18(1):88–104. doi: 10.20892/j.issn.2095-3941.2020.0096. PMID: 33628587; PMCID: PMC7877178.

44. Li J, Zhi X, Shen X, Chen C, Yuan L, Dong X, Zhu C, Yao L, Chen M. Depletion of UBE2C reduces ovarian cancer malignancy and reverses cisplatin resistance via downregulating CDK1. Biochem Biophys Res Commun. 2020 Mar 5;523(2):434–440. doi: 10.1016/j.bbrc.2019.12.058. Epub 2019 Dec 23. PMID: 31875843.

45. Martínez-Canales S, López de Rodas M, Nuncia-Cantarero M, Páez R, Amir E, Győrffy B, Pandiella A, Galán-Moya EM, Ocaña A. Functional transcriptomic annotation and protein-protein interaction analysis identify EZH2 and UBE2C as key upregulated proteins in ovarian cancer. Cancer Med. 2018 May;7(5):1896–1907. doi: 10.1002/cam4.1406. Epub 2018 Mar 25. PMID: 29575713; PMCID: PMC5943485.

46. Xiang C, Yan HC. Ubiquitin conjugating enzyme E2 C (UBE2C) may play a dual role involved in the progression of thyroid carcinoma. Cell Death Discov. 2022 Mar 24;8(1):130. doi: 10.1038/s41420-022-00935-4. PMID: 35332135; PMCID: PMC8948250.

47. Abreu RDS, Antunes D, Moreira ADS, Passetti F, Mendonça JB, de Araújo NS, Sassaro TF, Alberto AVP, Carrossini N, Fernandes PV, Costa MA, Guimarães ACR, Degrave WMS, Waghabi MC. Next Generation of Ovarian Cancer Detection Using Aptamers. Int J Mol Sci. 2023 Mar 28;24(7):6315. doi: 10.3390/ijms24076315. PMID: 37047289; PMCID: PMC10094455.

48. Vlasenkova R, Nurgalieva A, Akberova N, Bogdanov M, Kiyamova R. Characterization of SLC34A2 as a Potential Prognostic Marker of Oncological Diseases. Biomolecules. 2021 Dec 14;11(12):1878. doi: 10.3390/biom11121878. PMID: 34944522; PMCID: PMC8699446.

49. Wang Y, Zhang L, Luo X, et al. EPS8L1 promotes migration and metastasis of ovarian cancer by activating Rac1/MAPK signaling pathway via upregulating TIAM2. Authorea. December 16, 2022. DOI: 10.22541/au.167120661.17798120/v1.

50. Kim DK, Seo EJ, Choi EJ, Lee SI, Kwon YW, Jang IH, Kim SC, Kim KH, Suh DS, Seong-Jang K, Lee SC, Kim JH. Crucial role of HMGA1 in the self-renewal and drug resistance of ovarian cancer stem cells. Exp Mol Med. 2016 Aug 26;48(8):e255. doi: 10.1038/emm.2016.73. PMID: 27561949; PMCID: PMC5007643.

51. Masciullo V, Baldassarre G, Pentimalli F, Berlingieri MT, Boccia A, Chiappetta G, Palazzo J, Manfioletti G, Giancotti V, Viglietto G, Scambia G, Fusco A. HMGA1 protein over-expression is a frequent feature of epithelial ovarian carcinomas. Carcinogenesis. 2003 Jul;24(7):1191–8. doi: 10.1093/carcin/bgg075. Epub 2003 May 9. PMID: 12807722.

52. Ye C, Ren S, Sadula A, Guo X, Yuan M, Meng M, Li G, Zhang X, Yuan C. The expression characteristics of transmembrane protein genes in pancreatic ductal adenocarcinoma through comprehensive analysis of bulk and single-cell RNA sequence. Front Oncol. 2023 May 17;13:1047377. doi: 10.3389/fonc.2023.1047377. PMID: 37265785; PMCID: PMC10229874.

53. Zhao L, Li Y, Zhang Z, Zou J, Li J, Wei R, Guo Q, Zhu X, Chu C, Fu X, Yue J, Li X. Meta-analysis based gene expression profiling reveals functional genes in ovarian cancer. Biosci Rep. 2020 Nov 27;40(11):BSR20202911. doi: 10.1042/BSR20202911. PMID: 33135729; PMCID: PMC7677829.

54. Xi X, Cao T, Qian Y, Wang H, Ju S, Chen Y, Chen T, Yang J, Liang B, Hou S. CDC20 is a novel biomarker for improved clinical predictions in epithelial ovarian cancer. Am J Cancer Res. 2022 Jul 15;12(7):3303–3317. PMID: 35968331; PMCID: PMC9360218.

55. Li C, Li W, Zhang Y, Zhang X, Liu T, Zhang Y, Yang Y, Wang L, Pan H, Ji J, Wang C. Increased expression of antisense lncRNA SPINT1-AS1 predicts a poor prognosis in colorectal cancer and is negatively correlated with its sense transcript. Onco Targets Ther. 2018 Jul 10;11:3969–3978. doi: 10.2147/OTT.S163883. PMID: 30022840; PMCID: PMC6044340.

56. Feng H, Gu ZY, Li Q, Liu QH, Yang XY, Zhang JJ. Identification of significant genes with poor prognosis in ovarian cancer via bioinformatical analysis. J Ovarian Res. 2019 Apr 22;12(1):35. doi: 10.1186/s13048-019-0508-2. PMID: 31010415; PMCID: PMC6477749.

57. Sehovic E, Hadrovic A, Dogan S. Detection and analysis of stable and flexible genes towards a genome signature framework in cancer. Bioinformation. 2019 Nov 10;15(10):772–779. doi: 10.6026/97320630015772. PMID: 31831960; PMCID: PMC6900328.

58. Li N, Li H, Wang Y, Cao L, Zhan X. Quantitative proteomics revealed energy metabolism pathway alterations in human epithelial ovarian carcinoma and their regulation by the antiparasite drug ivermectin: data interpretation in the context of 3P medicine. EPMA J. 2020 Oct 10;11(4):661–694. doi: 10.1007/s13167-020-00224-z. PMID: 33240452; PMCID: PMC7680500.

59. He WP, Guo YY, Yang GP, Lai HL, Sun TT, Zhang ZW, Ouyang LL, Zheng Y, Tian LM, Li XH, You ZS, Xie D, Yang GF. CHD1L promotes EOC cell invasiveness and metastasis via the regulation of METAP2. Int J Med Sci. 2020 Aug 29;17(15):2387–2395. doi: 10.7150/ijms.48615. PMID: 32922205; PMCID: PMC7484650.

60. Soltan MA, Eldeen MA, Eid RA, Alyamani NM, Alqahtani LS, Albogami S, Jafri I, Park MN, Alsharif G, Fayad E, Mohamed G, Osman R, Kim B, Zaki MSA. A pan-cancer analysis reveals CHD1L as a prognostic and immunological biomarker in several human cancers. Front Mol Biosci. 2023 Mar 23;10:1017148. doi: 10.3389/fmolb.2023.1017148. PMID: 37033447; PMCID: PMC10076660.

61. He WP, Zhou J, Cai MY, Xiao XS, Liao YJ, Kung HF, Guan XY, Xie D, Yang GF. CHD1L protein is overexpressed in human ovarian carcinomas and is a novel predictive biomarker for patients survival. BMC Cancer. 2012 Sep 29;12:437. doi: 10.1186/1471-2407-12-437. PMID: 23020525; PMCID: PMC3551745.

62. Li Y, He LR, Gao Y, Zhou NN, Liu Y, Zhou XK, Liu JF, Guan XY, Ma NF, Xie D. CHD1L contributes to cisplatin resistance by upregulating the ABCB1-NF-κB axis in human non-small-cell lung cancer. Cell Death Dis. 2019 Feb 4;10(2):99. doi: 10.1038/s41419-019-1371-1. PMID: 30718500; PMCID: PMC6362241.

63. Saini U, Suarez AA, Naidu S, Wallbillich JJ, Bixel K, Wanner RA, Bice J, Kladney RD, Lester J, Karlan BY, Goodfellow PJ, Cohn DE, Selvendiran K. STAT3/PIAS3 Levels Serve as “Early Signature” Genes in the Development of High-Grade Serous Carcinoma from the Fallopian Tube. Cancer Res. 2018 Apr 1;78(7):1739–1750. doi: 10.1158/0008-5472.CAN-17-1671. Epub 2018 Jan 16. PMID: 29339537; PMCID: PMC5907493.

64. Vizeacoumar FJ, Arnold R, Vizeacoumar FS, Chandrashekhar M, Buzina A, Young JT, Kwan JH, Sayad A, Mero P, Lawo S, Tanaka H, Brown KR, Baryshnikova A, Mak AB, Fedyshyn Y, Wang Y, Brito GC, Kasimer D, Makhnevych T, Ketela T, Datti A, Babu M, Emili A, Pelletier L, Wrana J, Wainberg Z, Kim PM, Rottapel R, O’Brien CA, Andrews B, Boone C, Moffat J. A negative genetic interaction map in isogenic cancer cell lines reveals cancer cell vulnerabilities. Mol Syst Biol. 2013 Oct 8;9:696. doi: 10.1038/msb.2013.54. PMID: 24104479; PMCID: PMC3817404.

65. Zhang C, Xu C, Ma C, Zhang Q, Bu S, Zhang DL, Yu L, Wang H. TRPs in Ovarian Serous Cystadenocarcinoma: The Expression Patterns, Prognostic Roles, and Potential Therapeutic Targets. Front Mol Biosci. 2022 Jun 24;9:915409. doi: 10.3389/fmolb.2022.915409. PMID: 35813831; PMCID: PMC9263218.

66. Zhang Q, Wang X, Zhang X, Zhan J, Zhang B, Jia J, Chen J. TMEM14A aggravates the progression of human ovarian cancer cells by enhancing the activity of glycolysis. Exp Ther Med. 2022 Aug 5;24(4):614. doi: 10.3892/etm.2022.11551. PMID: 36160886; PMCID: PMC9468797.

67. Tian Y, Zhang J, Chen L, Zhang X. The expression and prognostic role of IMPDH2 in ovarian cancer. Ann Diagn Pathol. 2020 Jun;46:151511. doi: 10.1016/j.anndiagpath.2020.151511. Epub 2020 Mar 23. PMID: 32305001.

68. Onallah H, Mannully ST, Davidson B, Reich R. Exosome Secretion and Epithelial-Mesenchymal Transition in Ovarian Cancer Are Regulated by Phospholipase D. Int J Mol Sci. 2022 Oct 31;23(21):13286. doi: 10.3390/ijms232113286. PMID: 36362078; PMCID: PMC9658871.

69. Li H, Chen L, Tong X, Dai H, Shi T, Cheng X, Sun M, Chen K, Wei Q, Wang M. Functional genetic variants of CTNNBIP1 predict platinum treatment response of Chinese epithelial ovarian cancer patients. J Cancer. 2020 Sep 30;11(23):6850–6860. doi: 10.7150/jca.48218. PMID: 33123276; PMCID: PMC7592014.

70. Caetano MMM, Moreira GA, da Silva MR, Guimarães GR, Santos LO, Pacheco AA, Siqueira RP, Mendes FC, Marques Da Silva EA, Junior AS, Rangel Fietto JL, Saito Â, Boroni M, Bressan GC. Impaired expression of serine/arginine protein kinase 2 (SRPK2) affects melanoma progression. Front Genet. 2022 Sep 23;13:979735. doi: 10.3389/fgene.2022.979735. PMID: 36212152; PMCID: PMC9537589.

71. Jiang C, Yuan B, Hang B, Mao JH, Zou X, Wang P. FHOD1 is upregulated in gastric cancer and promotes the proliferation and invasion of gastric cancer cells. Oncol Lett. 2021 Oct;22(4):712. doi: 10.3892/ol.2021.12973. Epub 2021 Aug 5. PMID: 34457067; PMCID: PMC8358613.

72. Li Z, Guo Q, Zhang J, Fu Z, Wang Y, Wang T, Tang J. The RNA-Binding Motif Protein Family in Cancer: Friend or Foe? Front Oncol. 2021 Nov 4;11:757135. doi: 10.3389/fonc.2021.757135. PMID: 34804951; PMCID: PMC8600070.

73. Alexandrova E, Pecoraro G, Sellitto A, Melone V, Ferravante C, Rocco T, Guacci A, Giurato G, Nassa G, Rizzo F, Weisz A, Tarallo R. An Overview of Candidate Therapeutic Target Genes in Ovarian Cancer. Cancers (Basel). 2020 Jun 4;12(6):1470. doi: 10.3390/cancers12061470. PMID: 32512900; PMCID: PMC7352306.

74. El Khoury W, Nasr Z. Deregulation of ribosomal proteins in human cancers. Biosci Rep. 2021 Dec 22;41(12):BSR20211577. doi: 10.1042/BSR20211577. PMID: 34873618; PMCID: PMC8685657.

75. Wu F, Liu Y, Hu S, Lu C. Ribosomal protein L31 (RPL31) inhibits the proliferation and migration of gastric cancer cells. Heliyon. 2023 Jan 20;9(2):e13076. doi: 10.1016/j.heliyon.2023.e13076. PMID: 36816257; PMCID: PMC9936522.

76. Wang L, Sun L, Liu R, Mo H, Niu Y, Chen T, Wang Y, Han S, Tu K, Liu Q. Long non-coding RNA MAPKAPK5-AS1/PLAGL2/HIF-1α signaling loop promotes hepatocellular carcinoma progression. J Exp Clin Cancer Res. 2021 Feb 17;40(1):72. doi: 10.1186/s13046-021-01868-z. PMID: 33596983; PMCID: PMC7891009.

77. Sha Z, Zhou J, Wu Y, Zhang T, Li C, Meng Q, Musunuru PP, You F, Wu Y, Yu R, Gao S. BYSL Promotes Glioblastoma Cell Migration, Invasion, and Mesenchymal Transition Through the GSK-3β/β-Catenin Signaling Pathway. Front Oncol. 2020 Oct 15;10:565225. doi: 10.3389/fonc.2020.565225. PMID: 33178594; PMCID: PMC7593785.

78. Gao S, Sha Z, Zhou J, Wu Y, Song Y, Li C, Liu X, Zhang T, Yu R. BYSL contributes to tumor growth by cooperating with the mTORC2 complex in gliomas. Cancer Biol Med. 2021 Feb 15;18(1):88–104. doi: 10.20892/j.issn.2095-3941.2020.0096. PMID: 33628587; PMCID: PMC7877178.

79. Li Q, Ren CC, Chen YN, Yang L, Zhang F, Wang BJ, Zhu YH, Li FY, Yang J, Zhang ZA. A Risk Score Model Incorporating Three m6A RNA Methylation Regulators and a Related Network of miRNAs-m6A Regulators-m6A Target Genes to Predict the Prognosis of Patients With Ovarian Cancer. Front Cell Dev Biol. 2021 Sep 23;9:703969. doi: 10.3389/fcell.2021.703969. PMID: 34631700; PMCID: PMC8495156.

80. Kryczka J, Boncela J. Integrated Bioinformatics Analysis of the Hub Genes Involved in Irinotecan Resistance in Colorectal Cancer. Biomedicines. 2022 Jul 16;10(7):1720. doi: 10.3390/biomedicines10071720. PMID: 35885025; PMCID: PMC9312838.

81. Zhu QY, He ZM, Cao WM, Li B. The role of TSC2 in breast cancer: a literature review. Front Oncol. 2023 May 12;13:1188371. doi: 10.3389/fonc.2023.1188371. PMID: 37251941; PMCID: PMC10213421.

82. Cai J, Sun M, Hu B, Windle B, Ge X, Li G, Sun Y. Sorting Nexin 5 Controls Head and Neck Squamous Cell Carcinoma Progression by Modulating FBW7. J Cancer. 2019 Jun 2;10(13):2942–2952. doi: 10.7150/jca.31055. PMID: 31281471; PMCID: PMC6590026.

83. Lin C, Xin S, Huang X, Zhang F. PTPRA facilitates cancer growth and migration via the TNF-α-mediated PTPRA-NF-κB pathway in MCF-7 breast cancer cells. Oncol Lett. 2020 Nov;20(5):131. doi: 10.3892/ol.2020.11992. Epub 2020 Aug 20. PMID: 32934700; PMCID: PMC7471670.

84. Fierheller CT, Alenezi WM, Serruya C, Revil T, Amuzu S, Bedard K, Subramanian DN, Fewings E, Bruce JP, Prokopec S, Bouchard L, Provencher D, Foulkes WD, El Haffaf Z, Mes-Masson AM, Tischkowitz M, Campbell IG, Pugh TJ, Greenwood CMT, Ragoussis J, Tonin PN. Molecular Genetic Characteristics of FANCI, a Proposed New Ovarian Cancer Predisposing Gene. Genes (Basel). 2023 Jan 20;14(2):277. doi: 10.3390/genes14020277. PMID: 36833203; PMCID: PMC9956348.

85. Fierheller CT, Guitton-Sert L, Alenezi WM, Revil T, Oros KK, Gao Y, Bedard K, Arcand SL, Serruya C, Behl S, Meunier L, Fleury H, Fewings E, Subramanian DN, Nadaf J, Bruce JP, Bell R, Provencher D, Foulkes WD, El Haffaf Z, Mes-Masson AM, Majewski J, Pugh TJ, Tischkowitz M, James PA, Campbell IG, Greenwood CMT, Ragoussis J, Masson JY, Tonin PN. A functionally impaired missense variant identified in French Canadian families implicates FANCI as a candidate ovarian cancer-predisposing gene. Genome Med. 2021 Dec 3;13(1):186. doi: 10.1186/s13073-021-00998-5. PMID: 34861889; PMCID: PMC8642877.

86. Cordas Dos Santos DM, Eilers J, Sosa Vizcaino A, Orlova E, Zimmermann M, Stanulla M, Schrappe M, Börner K, Grimm D, Muckenthaler MU, Kulozik AE, Kunz JB. MAP3K7 is recurrently deleted in pediatric T-lymphoblastic leukemia and affects cell proliferation independently of NF-κB. BMC Cancer. 2018 Jun 18;18(1):663. doi: 10.1186/s12885-018-4525-0. PMID: 29914415; PMCID: PMC6006985.

87. Sakuma K, Sasaki E, Kimura K, Komori K, Shimizu Y, Yatabe Y, Aoki M. HNRNPLL stabilizes mRNA for DNA replication proteins and promotes cell cycle progression in colorectal cancer cells. Cancer Sci. 2018 Aug;109(8):2458–2468. doi: 10.1111/cas.13660. Epub 2018 Jul 16. PMID: 29869816; PMCID: PMC6113449.

88. Shi Y, Mo X, Hong S, Li T, Chen B, Chen G. Studying the Role and Molecular Mechanisms of MAP4K3 in Sorafenib Resistance of Hepatocellular Carcinoma. Biomed Res Int. 2020 Nov 5;2020:4965670. doi: 10.1155/2020/4965670. PMID: 33204699; PMCID: PMC7665914.

89. Muys BR, Sousa JF, Plaça JR, de Araújo LF, Sarshad AA, Anastasakis DG, Wang X, Li XL, de Molfetta GA, Ramão A, Lal A, Vidal DO, Hafner M, Silva WA. miR-450a Acts as a Tumor Suppressor in Ovarian Cancer by Regulating Energy Metabolism. Cancer Res. 2019 Jul 1;79(13):3294–3305. doi: 10.1158/0008-5472.CAN-19-0490. Epub 2019 May 17. PMID: 31101765; PMCID: PMC6606360.

90. Miśkiewicz J, Mielczarek-Palacz A, Gola JM. MicroRNAs as Potential Biomarkers in Gynecological Cancers. Biomedicines. 2023 Jun 13;11(6):1704. doi: 10.3390/biomedicines11061704. PMID: 37371799; PMCID: PMC10296063.

91. Zhao C, Li Y, Qiu C, Chen J, Wu H, Wang Q, Ma X, Song K, Kong B. Splicing Factor DDX23, Transcriptionally Activated by E2F1, Promotes Ovarian Cancer Progression by Regulating FOXM1. Front Oncol. 2021 Dec 13;11:749144. doi: 10.3389/fonc.2021.749144. PMID: 34966670; PMCID: PMC8710544.

92. Lisowska KM, Olbryt M, Dudaladava V, Pamuła-Piłat J, Kujawa K, Grzybowska E, Jarząb M, Student S, Rzepecka IK, Jarząb B, Kupryjańczyk J. Gene expression analysis in ovarian cancer - faults and hints from DNA microarray study. Front Oncol. 2014 Jan 28;4:6. doi: 10.3389/fonc.2014.00006. PMID: 24478986; PMCID: PMC3904181.

93. Zhang Z, Zhu Q. WD Repeat and HMG Box DNA Binding Protein 1: An Oncoprotein at the Hub of Tumorigenesis and a Novel Therapeutic Target. Int J Mol Sci. 2023 Aug 6;24(15):12494. doi: 10.3390/ijms241512494. PMID: 37569867; PMCID: PMC10420296.

94. Xian Q, Zhu D. The Involvement of WDHD1 in the Occurrence of Esophageal Cancer as a Downstream Target of PI3K/AKT Pathway. J Oncol. 2022 Apr 5;2022:5871188. doi: 10.1155/2022/5871188. PMID: 35422862; PMCID: PMC9005294.

95. Thomas H, Nasim MM, Sarraf CE, Alison MR, Love S, Lambert HE, Price P. Proliferating cell nuclear antigen (PCNA) immunostaining--a prognostic factor in ovarian cancer? Br J Cancer. 1995 Feb;71(2):357–62. doi: 10.1038/bjc.1995.72. PMID: 7841053; PMCID: PMC2033602.

96. Zhu X, You S, Du X, Song K, Lv T, Zhao H, Yao Q. LRRC superfamily expression in stromal cells predicts the clinical prognosis and platinum resistance of ovarian cancer. BMC Med Genomics. 2023 Jan 18;16(1):10. doi: 10.1186/s12920-023-01435-9. PMID: 36653841; PMCID: PMC9850808.

97. Wu J, Lu F, Yu B, Wang W, Ye X. The oncogenic role of SNRPB in human tumors: A pan-cancer analysis. Front Mol Biosci. 2022 Oct 6;9:994440. doi: 10.3389/fmolb.2022.994440. PMID: 36275630; PMCID: PMC9582665.

98. Dogan B, Gumusoglu E, Ulgen E, Sezerman OU, Gunel T. Integrated bioinformatics analysis of validated and circulating miRNAs in ovarian cancer. Genomics Inform. 2022 Jun;20(2):e20. doi: 10.5808/gi.21067. Epub 2022 Jun 30. PMID: 35794700; PMCID: PMC9299562.

99. Wagenbach M, Vicente JJ, Ovechkina Y, Domnitz S, Wordeman L. Functional characterization of MCAK/Kif2C cancer mutations using high-throughput microscopic analysis. Mol Biol Cell. 2020 Mar 19;31(7):580–588. doi: 10.1091/mbc.E19-09-0503. Epub 2019 Nov 20. PMID: 31746663; PMCID: PMC7202071.

100. Tong D, Volm T, Eberhardt E, Krainer M, Leodolter S, Kreienberg R, Zeillinger R. Rad52 gene mutations in breast/ovarian cancer families and sporadic ovarian carcinoma patients. Oncol Rep. 2003 Sep-Oct;10(5):1551–3. PMID: 12883740.

101. Diao Y, Li Y, Wang Z, Wang S, Li P, Kong B. SF3B4 promotes ovarian cancer progression by regulating alternative splicing of RAD52. Cell Death Dis. 2022 Feb 24;13(2):179. doi: 10.1038/s41419-022-04630-1. PMID: 35210412; PMCID: PMC8873359.

102. Challa S, Khulpateea BR, Nandu T, Camacho CV, Ryu KW, Chen H, Peng Y, Lea JS, Kraus WL. Ribosome ADP-ribosylation inhibits translation and maintains proteostasis in cancers. Cell. 2021 Aug 19;184(17):4531–4546.e26. doi: 10.1016/j.cell.2021.07.005. Epub 2021 Jul 26. PMID: 34314702; PMCID: PMC8380725.

103. Cao M, Deng Y, Deng Y, Wu J, Yang C, Wang Z, Hou Q, Fu H, Ren Z, Xia X, Li Y, Wang W, Xu H, Liao X, Shu Y. Characterization of immature ovarian teratomas through single-cell transcriptome. Front Immunol. 2023 Mar 3;14:1131814. doi: 10.3389/fimmu.2023.1131814. PMID: 36936909; PMCID: PMC10020330.

